# Dengue Viremia Kinetics and Effects on Platelet Count and Clinical Outcomes: An Analysis of 2340 Patients from Vietnam

**DOI:** 10.1101/2023.10.20.23297326

**Authors:** Nguyen Lam Vuong, Nguyen Than Ha Quyen, Nguyen Thi Hanh Tien, Duong Thi Hue Kien, Huynh Thi Le Duyen, Phung Khanh Lam, Dong Thi Hoai Tam, Tran Van Ngoc, Thomas Jaenisch, Cameron P. Simmons, Sophie Yacoub, Bridget A. Wills, Ronald B. Geskus

## Abstract

**Background:** Viremia is a critical factor in understanding the pathogenesis of dengue infection, but limited data exist on viremia kinetics. This study aimed to investigate the kinetics of viremia and its effects on subsequent platelet count, severe dengue, and plasma leakage.

**Methods:** We pooled data from three studies conducted in Vietnam between 2000 and 2016, involving 2340 dengue patients with daily viremia measurements and platelet counts after symptom onset. Viremia kinetics were assessed using a random effects model that accounted for left-censored data. The effects of viremia on subsequent platelet count and clinical outcomes were examined using a landmark approach with a random effects model and logistic regression model with generalized estimating equations, respectively. The rate of viremia decline was derived from the model of viremia kinetics. Its effect on the clinical outcomes was assessed by logistic regression models.

**Results:** Viremia levels rapidly decreased following symptom onset, with variations observed depending on the infecting serotype. DENV-1 exhibited the highest mean viremia levels during the first 5-6 days, while DENV-4 demonstrated the shortest clearance time. Higher viremia levels were associated with decreased subsequent platelet counts from day 6 onwards. Elevated viremia levels on each illness day increased the risk of developing severe dengue and plasma leakage. However, the effect size decreased with later illness days. A more rapid decline in viremia is associated with a reduced risk of the clinical outcomes.

**Conclusions:** This study provides comprehensive insights into viremia kinetics and its effect on subsequent platelet count and clinical outcomes in dengue patients. Our findings underscore the importance of measuring viremia levels during the early febrile phase for dengue studies and support the use of viremia kinetics as outcome for phase-2 dengue therapeutic trials.

## Introduction

Dengue is the most common arboviral infection worldwide and is found mainly in tropical and subtropical areas. The disease has been a growing threat for decades, and in 2019 the World Health Organization (WHO) ranked it among the top 10 threats to global health (World Health Organization, 2020). It is estimated that 105 million people are infected by dengue annually, of whom half develop symptoms (Cattarino, Rodriguez-Barraquer, Imai, Cummings, & Ferguson, 2020). Although most infections are asymptomatic or self-limiting, the small proportion of patients who develop complications can still overwhelm health systems in dengue-endemic countries during seasonal epidemics. Climate change, increasing global travel, and urbanization all serve to amplify the distribution of *Aedes aegypti*, the main vector responsible for dengue transmission (Whitehorn & Yacoub, 2019; Yacoub, Kotit, & Yacoub, 2011). Although dengue has been known for centuries, specific treatment is still to be found, and the two licensed vaccines are of limited benefit (Kariyawasam, Lachman, Mansuri, Chakrabarti, & Boggild, 2023; Redoni et al., 2020).

Exploring the variation in viremia levels could help improve our understanding of disease pathogenesis and potentially facilitate assessment of new vaccines and treatments for dengue. For example, knowing the natural kinetics of dengue viremia may help in selection of patient groups or timing of antiviral use in therapeutic intervention trials. Dengue viremia kinetics has been investigated in several studies (Ben-Shachar, Schmidler, & Koelle, 2016; Clapham et al., 2016; Clapham, Tricou, Van Vinh Chau, Simmons, & Ferguson, 2014; Duyen et al., 2011; Matangkasombut et al., 2020; Nguyet et al., 2013; Simmons, Chau, et al., 2007; Tricou, Minh, Farrar, Tran, & Simmons, 2011). The main findings are: (i) viremia rapidly decreases after symptom onset; (ii) dengue virus (DENV)-1 infection gives higher viremia level than DENV-2 and 3; and (iii) primary infection gives higher viremia than secondary infection in DENV-1. However, these studies had limited sample sizes, particularly for DENV-4 serotype, and did not provide a full account of viremia kinetics. Three studies constructed mechanistic mathematical models and focused on investigating the role of the host immune response in controlling viremia (Ben-Shachar et al., 2016; Clapham et al., 2016; Clapham et al., 2014). Another study assessed viremia in infants aged under 18 months (Simmons, Chau, et al., 2007); the findings cannot be generalized to older children or adults. Others focused on peak viremia level after symptom onset, time to viral clearance and/or the probability of detecting virus on each illness day (Duyen et al., 2011; Matangkasombut et al., 2020; Nguyet et al., 2013; Tricou et al., 2011). In most analyses, viremia levels below the limit of detection were simply set as zero, which may not be appropriate.

Quantifying the effect of viremia level on clinical outcomes is important to understand the mechanisms leading to severe disease. We and others have previously found that higher viremia levels are associated with more severe dengue (Duyen et al., 2011; Endy et al., 2004; Morsy et al., 2020; Tang et al., 2010; Tricou et al., 2011; Vaughn et al., 2000; Vuong et al., 2021), but the effect size was not strong (Vuong et al., 2021). However, all analyses were based on a single viremia value per individual. Investigating how individual viremia levels at different times after symptom onset affect the risk of severe outcome potentially provides new insights into pathogenic mechanisms. Another important question is whether a more rapid decline in viremia is associated with a lower risk of more severe clinical outcomes. One study has shown a slower rate of viral clearance being associated with more severe outcomes (Wang et al., 2006), while others have shown no association (Fox et al., 2011; Vaughn et al., 2000). This lack of conclusive evidence underscores the need for further investigation. In addition, the relationship between viremia kinetics and platelet count kinetics – an important hematological indicator – has yet to be investigated.

This study aims to improve our understanding of viremia kinetics, how it is associated with patient characteristics and virus serotype, and how it impacts disease severity, using a dataset derived from several thousand individuals with symptomatic dengue. First, we fitted a model for individual viremia kinetics that treats values below the detection limit as censored data, and assessed whether viremia kinetics differed by age, sex, serotype, or immune status. Second, we modeled platelet count kinetics and explored dependence on viremia level. We investigated the potential existence of a lagged effect of viremia, by analyzing platelet counts measured on and following the day of viremia assessment. Third, we modeled the effect of viremia levels measured at different days after symptom onset and the effect of the rate of viremia decline on development of two clinical outcomes, plasma leakage and severe dengue.

## Methods

### Study population

We used data from three prospective observational studies performed as part of the longstanding collaboration between the Oxford University Clinical Research Unit (OUCRU) and the Hospital for Tropical Diseases (HTD) in Ho Chi Minh City, Vietnam. Protocol synopses for the three studies are presented in **Appendix 1**. Briefly, studies A and B simultaneously ran from 2000 to 2009 and enrolled children aged between 5 and 15 years. Study A included children with a febrile illness presenting to community outpatient clinics, while study B included children admitted to HTD with a febrile illness suspected to be dengue. Some patients from study A were subsequently enrolled in study B after admission to hospital. Study C ran from 2011 to 2016 and enrolled adults and children presenting with possible dengue within three days of fever onset (IDAMS study, NCT01550016) (Jaenisch et al., 2016). Clinical evaluation and blood sampling were performed daily for all three studies. All studies were approved by the Oxford Tropical Research Ethics Committee and the Institutional Ethics Committee.

We identified 3527 laboratory-confirmed dengue patients. Among them, 2340 had relevant data on viremia kinetics and were included in this analysis (**Figure 1**). Details of dengue diagnostics are in **Appendix 2**.

**Figure 1.**
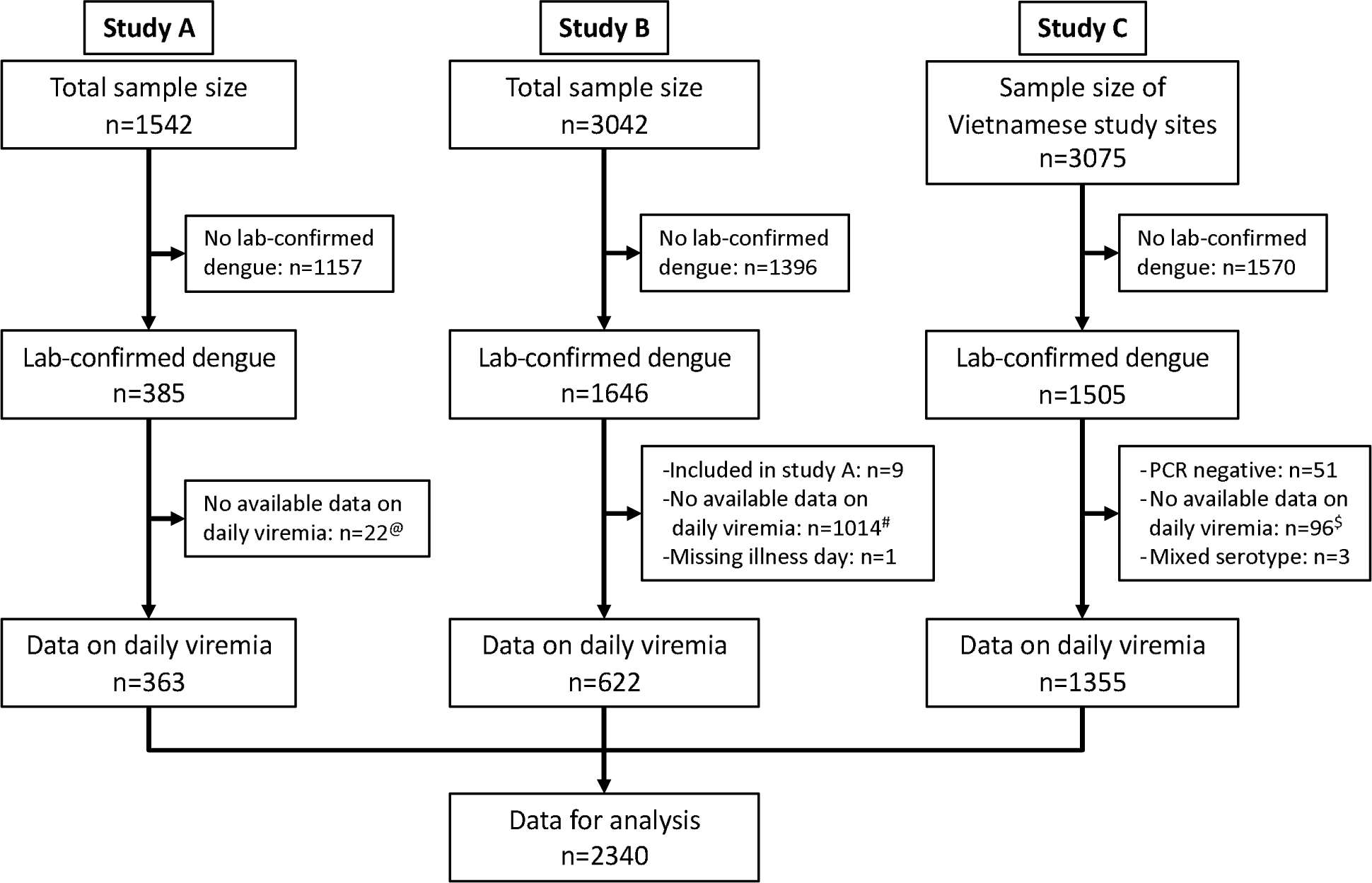
Flowchart illustrating the process of patient selection. ^@^Among 385 lab-confirmed dengue patients in study A, 31 individuals were later included in study B upon hospitalization. Of these, 9 had viremia measurements available in both studies and were consequently analysed in study A. The remaining 22 lacked viremia data in study A but had measurements in study B, leading to their inclusion in study B in the analysis. ^#^Although study B spanned from 2000 to 2009, daily viremia levels were only measured between 2006 and 2008. ^$^In study C, one study site in Vietnam did not have daily viremia measurements. Overall, a random sample of 2340 from 3527 laboratory-confirmed dengue patients had data on viremia kinetics. PCR, polymerase chain reaction.

### Plasma viremia measurement, dengue diagnostics, and clinical endpoints (details in Appendix 2)

Plasma viremia levels were measured by reverse transcription polymerase chain reaction (RT-PCR), which was an internally controlled, serotype-specific, real-time, two-step assay in studies A and B (Simmons, Popper, et al., 2007), and a one-step procedure using a validated assay in study C (Hue et al., 2011). There was no formal validation of the detection limit for viremia for the two-step PCR used in studies A and B; we set it at 1000 copies/ml. In the one-step PCR used in study C, the detection limit was 300 copies/ml for DENV-1 and DENV-3, 60 copies/ml for DENV-2, and 600 copies/ml for DENV-4 (Hue et al., 2011).

Patients were classified into probable primary (i.e., the first) or probable secondary (i.e., a second or subsequent) infection based on IgG results on paired samples. A probable primary infection was defined by two negative/equivocal IgG results on separate samples taken at least two days apart within the first ten days of symptom onset, with at least one sample during the convalescent phase (days 6-10). A probable secondary infection was defined by at least one positive IgG result during the first ten days. Cases without time-appropriate IgG results were classified as indeterminate.

For the clinical endpoints we selected severe dengue and plasma leakage. The definitions are based on the WHO 2009 guidelines (World Health Organization, 2009) and standard endpoint definitions for dengue trials (Tomashek et al., 2018). Severe dengue was defined as severe plasma leakage, severe bleeding, and/or severe organ impairment. Plasma leakage included moderate and severe leakage.

### Statistical analysis (details in Appendix 3)

In **Appendix 3-figure 1** we show a directed acyclic graph (DAG) to display assumptions about the causal relationships between variables over illness day. Illness day is the number of days after symptom onset, where day 1 is the day of symptom onset. Potential confounders that needed to be corrected for were age, sex, DENV serotype, and primary/secondary immune status. In all analyses, we used a log-10 transformation for viremia levels.

We modeled viremia kinetics over time and explored dependence on age, sex, serotype, immune status and PCR method using a linear mixed-effects regression model, allowing for left censored values in the outcome variable. In the fixed effect, we allowed for interactions between serotype and immune status, and between illness day and all other covariates. Nonlinear trends by age and illness day were investigated using splines with three and four knots, respectively. We included a random effect with an intercept and splines for illness day with four knots to model the intra-person correlation. Due to the presence of a left-skewed distribution in log-10 viremia (**Appendix 4-figure 1**), a sensitivity analysis was conducted using the 10th-root transformation.

Platelet counts were transformed using a fourth-root transformation since the distribution was right-skewed (**Appendix 4-figure 2**). The effect of viremia on subsequent platelet count was investigated using the landmark approach (van Houwelingen & Putter, 2011). For each illness day from 1 to 7, a landmark dataset was created, which included viremia on that day, platelet count from that day to day 10, and the time-fixed variables (age, sex, serotype, immune status, and PCR method). For the supermodel that combined all landmark datasets, we used a linear mixed effects model including viremia, age, sex, serotype, immune status, PCR method, illness day and landmark. In the fixed effect, we also included four-way interactions between viremia, serotype, PCR method and a) and immune status, b) illness day, and c) landmark day. Splines with three knots were used to allow for nonlinear trends in log-10 viremia, age, illness day, and landmark day. We included a random effect with an intercept by the combination of individuals and landmark, and splines for illness day with three knots.

The effect of viremia on the clinical endpoints was investigated using a logistic regression model using the landmark approach (van Houwelingen & Putter, 2011). For each illness day from 1 to 7, a landmark dataset was created, which included viremia on that day, occurrence of the endpoint on or after that day, and the time-fixed variables similar to above. All individuals who had already experienced the relevant endpoint before that day were excluded. We fitted a logistic regression model for each landmark dataset and then fitted a supermodel that combined all the datasets. For the supermodel we used generalized estimating equations (GEE) to account for repeated inclusion of individuals at multiple landmarks. We included viremia, age, sex, serotype, immune status, study, and landmark day, and allowed for the four-way interaction between viremia, serotype, immune status and landmark day, and two-way interactions between viremia and all other covariates. Splines were used to allow for nonlinear trends in log-10 viremia (three knots), age (three knots), and landmark (three knots). Since plasma leakage had missing data, we performed an analysis with imputed data using multiple imputation by chained equations (MICE). The imputation was done in our previous study (Vuong et al., 2021).

To calculate the rate of viremia decline for each patient, we assumed a linear decline in log-10 viremia over time. We modified our viremia kinetics model by using a linear term for illness day rather than the original nonlinear terms, both in the fixed and in the random effect. For each patient, we then calculated fitted values using this model. The rate of viremia decline was defined as the daily difference in log-10 viremia. We assessed the impact of the decline rate on the clinical endpoints using logistic regression models. We did not use the landmark approach for this analysis since the decline rate was assumed to be constant over time. Covariates included the decline rate, serotype, immune status, and PCR method. Due to limited number of severe dengue cases, its model did not include any interaction terms. The plasma leakage model incorporated interactions between the decline rate and all other covariates.

In the analyses for platelet count and clinical outcomes, undetectable viremia levels were set as the specific detection limits, and a binary dummy variable (yes/no) was created to indicate whether the viremia value was undetectable. Since the individual spline terms are hard to interpret and the large data set makes most differences statistically significant, results are primarily reported using plots of predicted values. All analyses were done using the statistical software R version 4.1.0 (R Core Team, 2021) with ‘MCMCglmm’ (Hadfield, 2010), ‘lme4’ (Bates, Maechler, Bolker, & Walker, 2015), ‘geepack’ (Halekoh, Højsgaard, & Yan, 2006), and ‘rms’ (Harrell Jr, 2021) packages for the models.

## Results

Baseline characteristics and outcomes are summarized in **Table 1**. Three quarters of the participants were children, and male sex predominated (60%). Most patients (85%) were enrolled on illness day 2 or 3. All four serotypes were included, but DENV-1 was the most prevalent (54%). Most infections (69%) were classified as probable secondary infection, while in 11% immune status could not be determined. There were 353 patients (15%) with plasma leakage and 65 patients (3%) with severe dengue. In **Appendix 4-table 1** we summarize plasma leakage and severe dengue by serotype and immune status. DENV-2 had the highest proportion of these two outcomes. Patients with probable secondary infection were more likely to experience these outcomes than those with probable primary infection or indeterminate immune status, regardless of the infecting serotype.

**Table 1.** Baseline characteristics and clinical outcomes.

|  | N<sup>a</sup> | All patients<br>(N=2340) | Study A<br>(N=363) | Study B<br>(N=622) | Study C<br>(N=1355) |
| --- | --- | --- | --- | --- | --- |
| Age, years | 2340 | 13 (10; 18) | 12 (9; 14) | 12 (10; 13) | 16 (10; 25) |
| Sex male | 2340 | 1403 (60) | 195 (54) | 412 (66) | 796 (59) |
| Illness day at enrolment | 2340 |  |  |  |  |
| 1 |  | 310 (13) | 59 (16) | 7 (1) | 244 (18) |
| 2 |  | 848 (36) | 150 (41) | 155 (25) | 543 (40) |
| 3 |  | 1137 (49) | 114 (31) | 455 (73) | 568 (42) |
| 4 |  | 45 (2) | 40 (11) | 5 (1) | 0 (0) |
| Serotype | 2340 |  |  |  |  |
| DENV-1 |  | 1264 (54) | 233 (64) | 410 (66) | 621 (46) |
| DENV-2 |  | 373 (16) | 48 (13) | 130 (21) | 195 (14) |
| DENV-3 |  | 252 (11) | 80 (22) | 82 (13) | 90 (7) |
| DENV-4 |  | 451 (19) | 2 (1) | 0 (0) | 449 (33) |
| Immune status | 2340 |  |  |  |  |
| Probable primary |  | 474 (20) | 134 (37) | 124 (20) | 216 (16) |
| Probable secondary |  | 1619 (69) | 219 (60) | 464 (75) | 936 (69) |
| Indeterminate |  | 247 (11) | 10 (3) | 34 (5) | 203 (15) |
| Plasma leakage | 2288 | 353 (15) | 43 (13) | 177 (29) | 133 (10) |
| Missing |  | 52 | 39 | 13 | 0 |
| Illness day of plasma leakage | 327 |  |  |  |  |
| 3 |  | 2 (1) | 0 (0) | 0 (0) | 2 (2) |
| 4 |  | 71 (22) | 12 (30) | 35 (23) | 24 (18) |
| 5 |  | 107 (33) | 12 (30) | 50 (32) | 45 (34) |
| 6 |  | 105 (32) | 12 (30) | 49 (32) | 44 (33) |
| 7 |  | 42 (13) | 4 (10) | 20 (13) | 18 (14) |
| Missing |  | 26 | 3 | 23 | 0 |
| Severe dengue | 2340 | 65 (3) | 6 (2) | 40 (6) | 19 (1) |
| Illness day of severe dengue | 65 |  |  |  |  |
| 3 |  | 2 (3) | 0 (0) | 2 (5) | 0 (0) |
| 4 |  | 13 (20) | 0 (0) | 12 (30) | 1 (5) |
| 5 |  | 24 (37) | 2 (33) | 16 (40) | 6 (32) |
| 6 |  | 20 (31) | 4 (67) | 7 (18) | 9 (47) |
| 7 |  | 6 (9) | 0 (0) | 3 (8) | 3 (16) |

<sup>a</sup> N represents the number of patients with available data (i.e., without missing data).
Summary statistics are number of patients (%) or median (25<sup>th</sup>; 75<sup>th</sup> percentiles).
DENV, dengue virus

### Viremia kinetics and the relationship with clinical characteristics

Individual viremia trajectories are shown in **Figure 2**. Most individuals had a decreasing trend from day 1 or 2 onwards. However, values higher than the baseline value were observed up to day 7 (**Appendix 4-figure 3**). The decreasing trend was consistent across all combinations of serotype and immune status (**Figure 2**).

**Figure 2.**
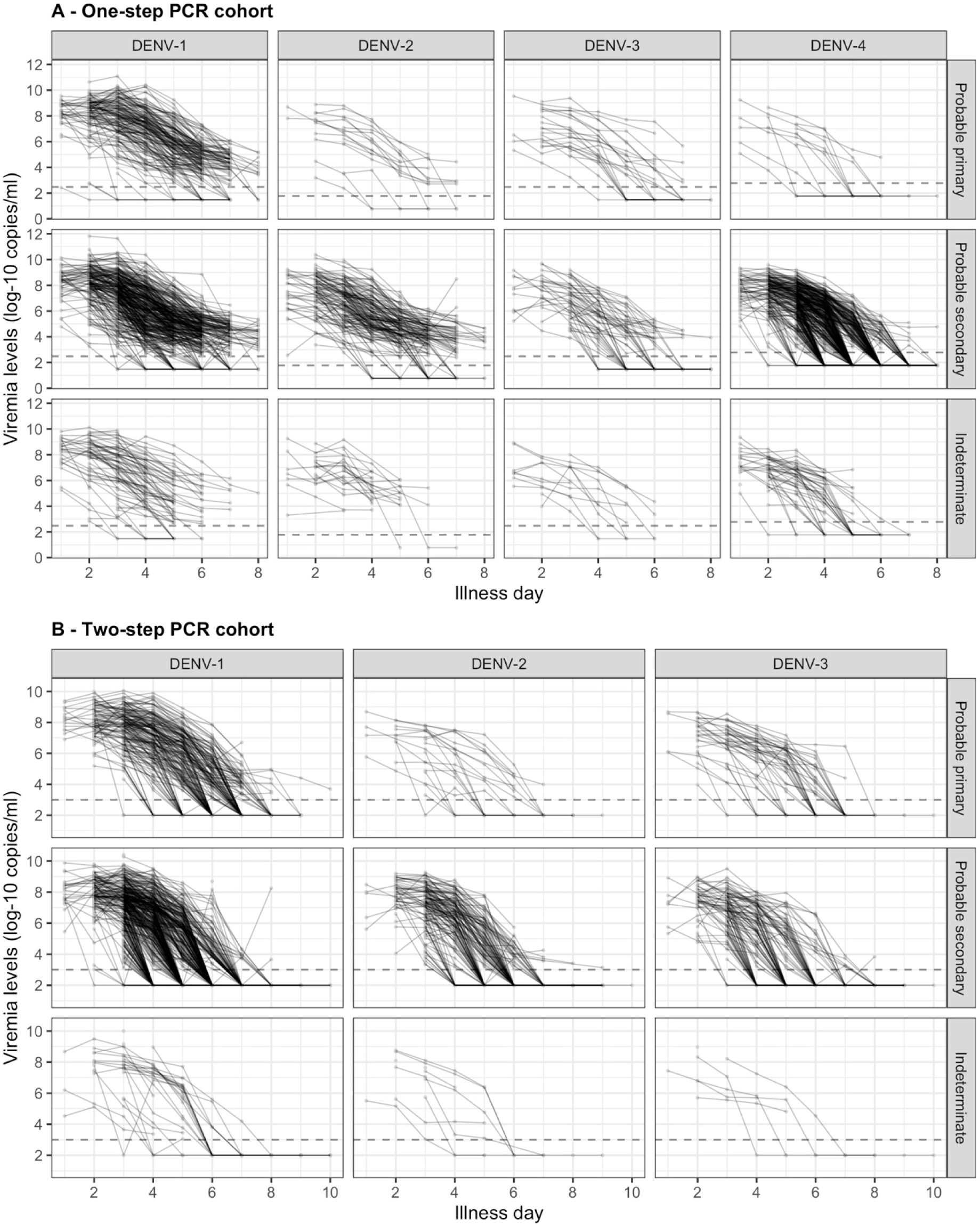
Individual trajectories of measured viremia levels. The dots represent the measured viremia levels and are connected by lines for each individual patient. The dashed lines indicate the detection limits, which are 300 copies/ml for DENV-1 and DENV-3, 60 copies/ml for DENV-2, and 600 copies/ml for DENV-4 in the one-step PCR cohort, and 1000 copies/ml in the two-step PCR cohort. 57 measured values that are lower than 1000 copies/ml in the two-step PCR cohort are considered as below the detection limit. Values below the detection limit are visually represented as 1/10 of the corresponding detection limit (on the original scale). DENV, dengue virus; PCR, polymerase chain reaction.

In **Figure 3** we present the fitted values based on the model for viremia for the one-step PCR cohort. The mean viremia trajectory differed by serotype. DENV-1 gave the highest viremia levels, with DENV-2 in secondary infection giving similar levels from day 5 onwards. DENV-2, DENV-3 and DENV-4 had similar viremia levels during the first three days. However, viremia decreased rapidly in DENV-4, reaching undetectable levels in the shortest time, while DENV-2 showed the slowest decline. These differences between serotypes were more pronounced in case of probable primary infection (**Figure 3-A**). In terms of immune status, probable primary infection showed higher viremia levels compared to probable secondary infection in DENV-1 from day 3 onwards. The disparity between probable primary and probable secondary infection was less pronounced in the other serotypes (**Figure 3-B**). Mean viremia levels were comparable between females and males (**Figure 3-C**), as well as across age (**Figure 3-D**). The one-step PCR method resulted in longer viremia compared to the two-step PCR (**Appendix 5-figure**). **1** The sensitivity analysis using the 10th-root transformation of viremia gave results similar to the main analysis using log-10 viremia (**Appendix 5-figure 2**).

**Figure 3.**
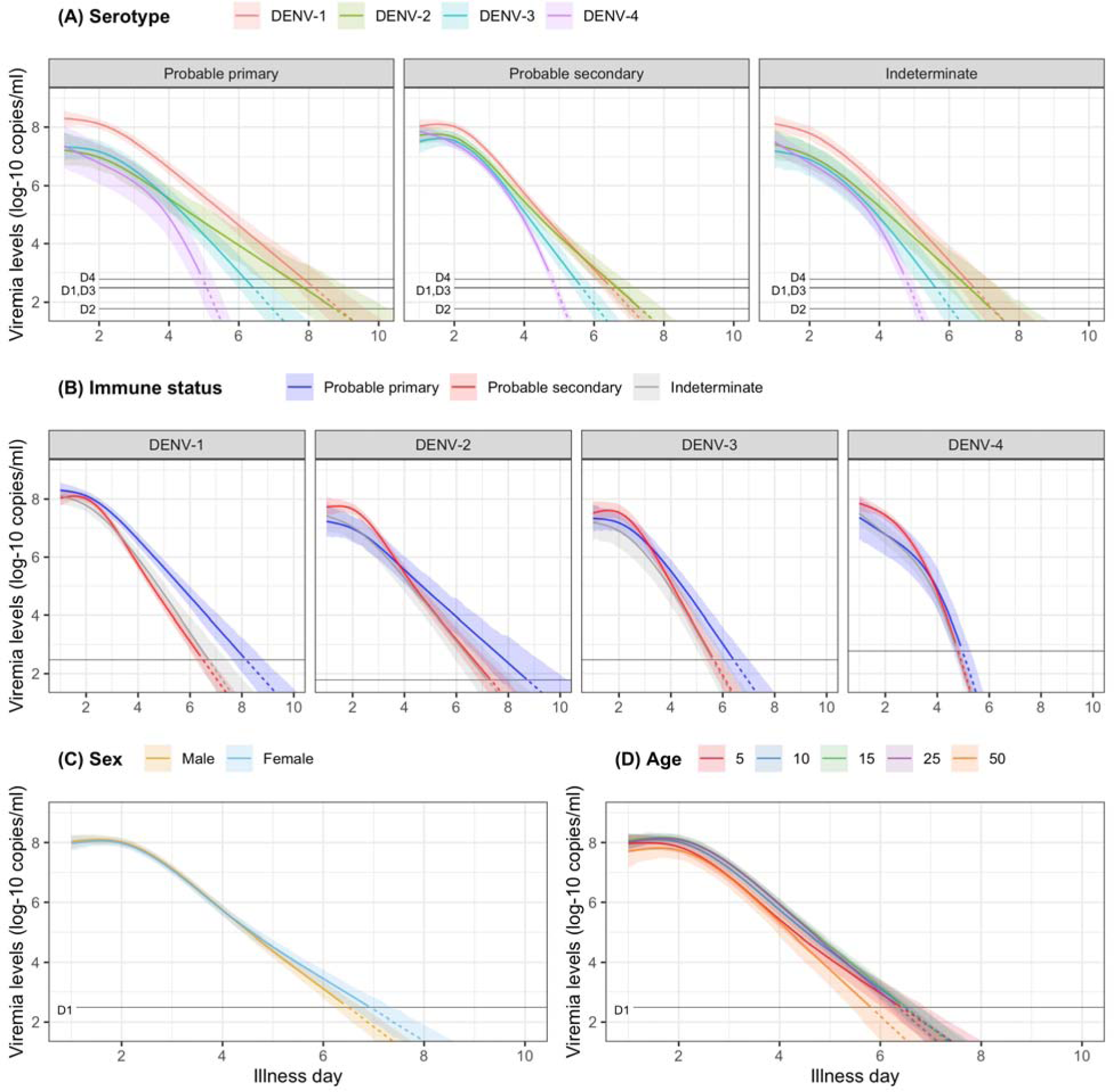
Fitted trends in mean viremia levels. The colored lines represent the estimated mean viremia levels, and the colored shaded regions represent the corresponding 95% credible intervals. The horizontal lines represent the detection limits (D1, D2, D3, and D4 denote DENV-1, DENV-2, DENV-3, and DENV-4, respectively). Dashed lines indicate fitted viremia levels that are below the detection limit. Viremia levels are shown for age of 10 years, male sex, serotype DENV-1, probable secondary infection, and using the one-step PCR. DENV, dengue virus; PCR, polymerase chain reaction.

### Effect of viremia on subsequent platelet count

Figure 4 shows the mean platelet counts by viremia levels based on the supermodel for the one-step PCR method. There was minimal impact of viremia on days 1-5 platelet count. However, higher viremia levels gave decreased subsequent platelet counts from day 6 onwards, for all serotypes. The strength of this effect increased with later illness day and was more pronounced in DENV-1 and DENV-2 compared to DENV-3 and DENV-4. The effect of viremia on subsequent platelet count remained consistent across subgroups of immune status, sex, and age. In the two-step PCR cohort, the effect of viremia on platelet count was less pronounced compared to the one-step PCR cohort, although the overall trends were similar (**Appendix 6-figure 1**).

**Figure 4.**
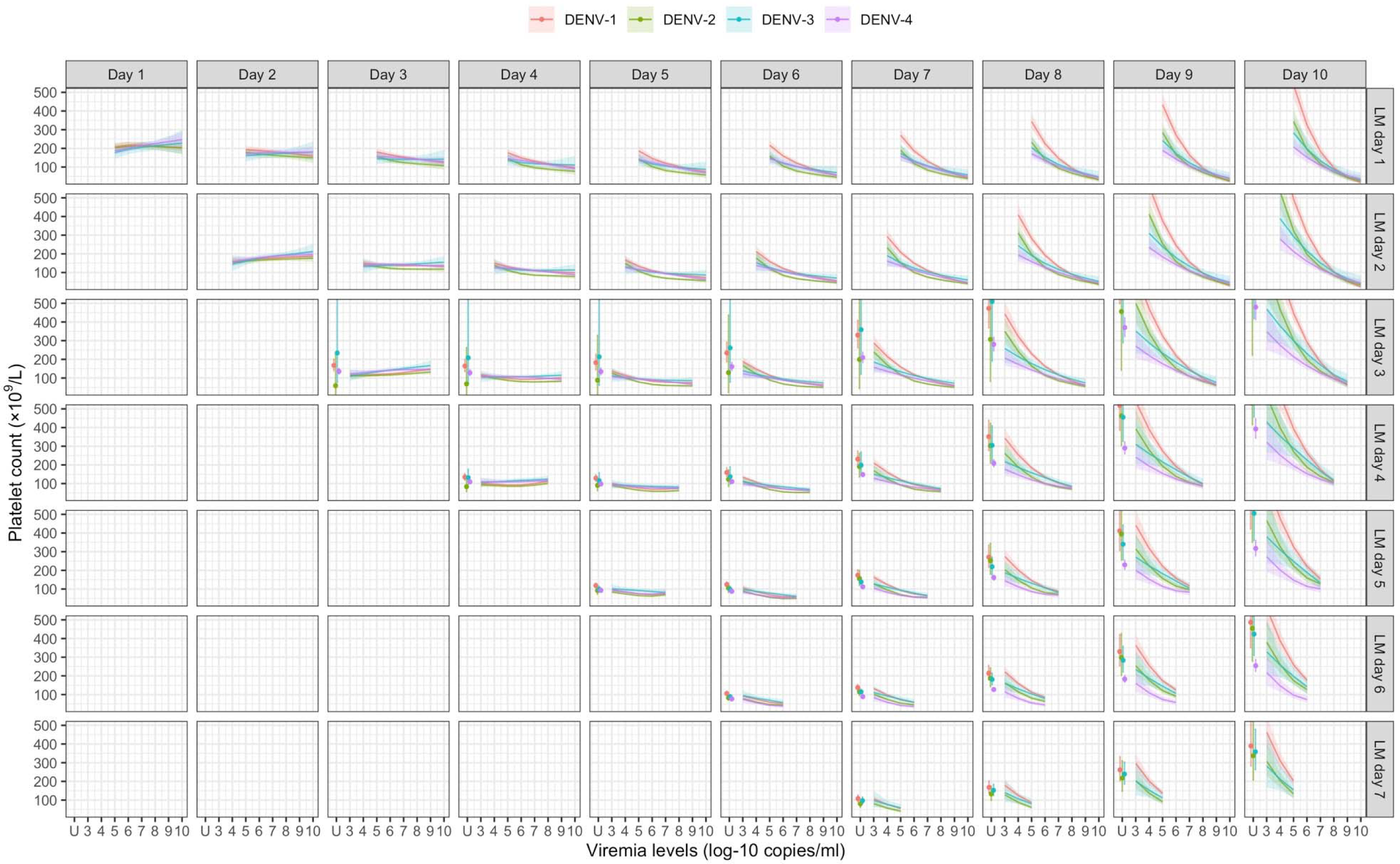
Fitted trends in mean platelet counts according to viremia levels – results from the supermodel. The colored lines or dots represent the estimated mean platelet counts. The colored shaded regions and whiskers indicate the corresponding 95% confidence intervals. Each row represents the effect of viremia on a specific day to platelet count from that day to day 10. No fitted trends are made for DENV-4 in LM day 7 since viremia was undetectable in almost all DENV-4 cases from day 7 onwards. The mean platelet counts are shown for age of 10 years, male sex, probable secondary infection, and using the one-step PCR. DENV, dengue virus; LM, landmark; PCR, polymerase chain reaction; U, under the limit of detection.

### Effect of viremia on clinical outcomes

Higher viremia levels increased the risk of severe dengue and plasma leakage for each of the serotypes (Figure 5). The effect of viremia on both endpoints decreased with later landmark day of viremia measurement, particularly for severe dengue (**Appendix 7-table 1**). However, these trends were not clear in the results obtained from the simple logistic regression models at each landmark time point (**Appendix 7-figure 1**). This trend remained consistent across subgroups of sex, age, and study (**Appendix 7-figure 2**). Furthermore, the results were consistent between the analyses with and without imputation (**Appendix 7-figure 3**).

**Figure 5.**
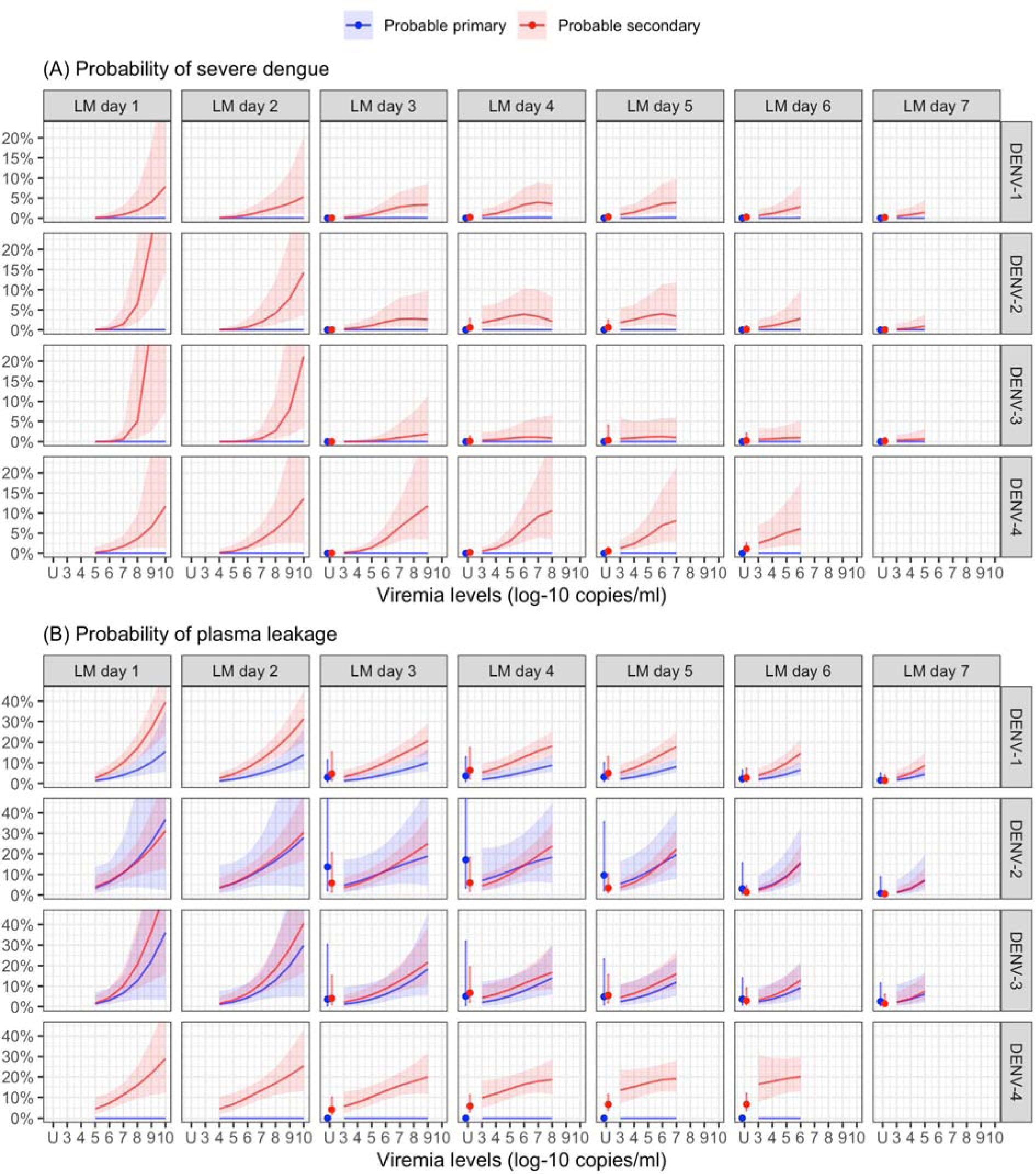
Probability of occurrence of the two clinical endpoints according to viremia levels – results from the supermodel. The colored lines or dots represent the probability of the endpoints. The colored shaded regions and whiskers indicate the corresponding 95% confidence intervals. Each column represents the effect of viremia on a specific day. No fitted trends are made for DENV-4 in LM day 7 since viremia was undetectable in almost all DENV-4 cases from day 7 onwards. Note that due to the limited number of severe dengue in primary infections (only 1 case in DENV-1) and plasma leakage in primary DENV-4 (see Appendix 4-table 1), the estimated probability of having these outcomes is nearly zero across all viremia levels within these subgroups. The probabilities are shown for age of 10 years, male sex, probable secondary infection, and from Study C. DENV, dengue virus, LM, landmark; PCR, polymerase chain reaction; U, under the limit of detection.

The supermodel revealed that older individuals had a relatively lower risk of developing severe dengue, but the trend was not significant. Males exhibited a slightly higher risk of severe dengue compared to females. The effect of serotype on the two endpoints was dependent on immune status. Individuals with probable secondary infection had a higher risk of experiencing both endpoints compared to those with probable primary infection (**Appendix 7-figure 4**).

From the model of viremia kinetics with a linear trend of illness day, the individual rate of viremia decline ranged from 0.58 to 2.01 log-10 copies/ml/day, with a median of 1.39 log-10 copies/ml/day. The logistic regression model demonstrated that a faster decline in viremia reduced the risk of severe dengue and plasma leakage (Figure 6). The effect on plasma leakage was stronger in the probable secondary infection group (**Appendix 7-table 2**).

**Figure 6.**
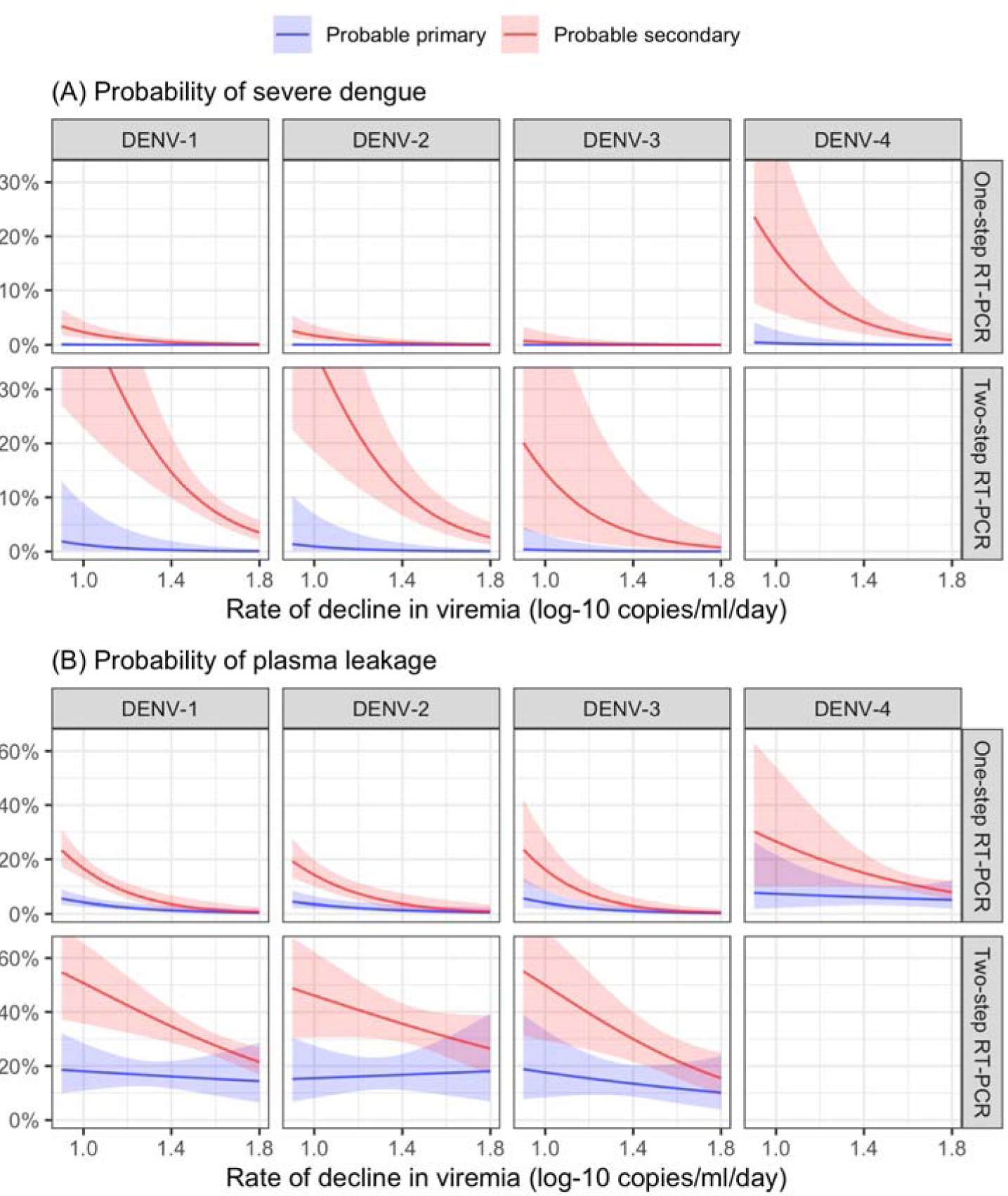
Probability of occurrence of the two clinical endpoints according to the rate of decline in viremia. The colored lines represent the probability of the endpoints. The colored shaded regions indicate the corresponding 95% confidence intervals.

## Discussion

In this study, we conducted a comprehensive analysis using a large dataset of 2340 viremic dengue patients to examine viremia kinetics from first presentation, its association with various virus and patient characteristics, and its impact on subsequent platelet count and disease severity. As others have described, we found that plasma viremia declines rapidly following the onset of symptoms, with the kinetics primarily influenced by the specific infecting serotype. Higher viremia levels were associated with a decrease in subsequent platelet count from day 6 onwards. Elevated viremia levels were found to increase the risk of developing severe dengue and plasma leakage, with the effect becoming weaker at later days. The effects of viremia on platelet count and clinical outcomes did not differ much by different subgroups of serotype, immune status, age, and sex. Moreover, a faster decline in viremia was associated with a reduced risk of the more severe clinical outcomes.

There is a limited number of published papers that studied the individual trajectory of dengue viremia. Most used data from Vietnam (Ben-Shachar et al., 2016; Clapham et al., 2016; Clapham et al., 2014; Duyen et al., 2011; Nguyet et al., 2013; Simmons, Chau, et al., 2007; Tricou et al., 2011), while one was from Thailand (Matangkasombut et al., 2020). Our study has provided further evidence supporting previous findings. We confirmed that (i) viremia rapidly decreases following the onset of symptoms, (ii) DENV-1 exhibits higher viremia levels compared to DENV-2 and DENV-3, and (iii) primary infection is associated with higher and more prolonged detectable viremia than secondary infection in DENV-1, while this pattern was not consistent across other serotypes. We have added viremia kinetics of DENV-4, and showed that viremia levels declined more rapidly than the other serotypes. Furthermore, our study demonstrated that the new PCR test has the ability to detect plasma viremia for a longer period compared to the older test. This may be attributed to the lower detection limits of the new test. Explaining the differences in viremia kinetics between serotypes remains challenging, as the molecular factors of the virus that influence plasma viremia levels are still unknown. It is also important to note that our study focused on the time since symptom onset rather than the time since infection. We cannot rule out that some of the observed differences are explained by a difference in time from infection to symptom onset.

Our study suggests that higher viremia levels on any day before day 6 are associated with reduced platelet count on days 6-8, typically corresponding to the nadir of platelet count. Additionally, it is possible that viremia in the initial four days after symptom onset could affect platelet count with a delay of 3-4 days. The direct involvement of the dengue virus in triggering platelet activation and apoptosis can contribute to the development of thrombocytopenia. Thrombocytopenia in dengue infection is thought to occur through two mechanisms: bone marrow suppression, which reduces thrombopoiesis, and increased peripheral platelet clearance (Quirino-Teixeira, Andrade, Pinheiro, Rozini, & Hottz, 2021). Various processes contribute to these mechanisms, including platelet-leukocyte and platelet-endothelial cell interactions, phagocytosis, complement-mediated lysis, aggregation, and clot formation. Additionally, several host immune response factors are involved in platelet activation (Balakrishna Pillai, Chu, Mariappan, & JeanPierre, 2021). In our analysis, we considered interactions between viremia level and both serotype as well as immune status. Interestingly, we observed that the effect of viremia level on subsequent platelet count did not differ much by serotype and immune status.

Higher plasma viremia levels increase the risk of worse clinical outcomes, as demonstrated in our previous study that only utilised viremia at enrollment during the febrile phase (Vuong et al., 2021). The diminishing impact of viremia on the two endpoints on later illness days may be attributed to the heightened immune responses that are likely triggered by higher viremia levels, and the resulting complex interplay between these factors that underlies progression to severe dengue. The effects of viremia, age, serotype, immune status, and illness day on the clinical endpoints in this study are consistent with those observed in the previous study. The similarity between these two analyses, along with the weaker effect of viremia at later days, suggests that viremia levels around the time of symptom onset could serve as a reliable predictor of dengue severity. It suggests that the early febrile phase, specifically illness day 1-3, is the critical period of measuring viremia levels in clinical practice and dengue studies. Secondary infection remains a substantial risk factor for more severe outcomes, while viremia kinetics are not influenced much by immune status. These findings suggest that immune status and viremia may be independent predictors of clinical outcomes, following distinct pathways.

The landmark approach enabled us to investigate the effect of viremia on platelet count and the clinical endpoints at each illness day, ranging from day 1 to 7, while ensuring that the viremia measurements preceded the occurrence of the outcomes. The supermodel allows for investigation of trends of the effects, while gaining power because we assume the effects of some fixed factors (e.g., age and sex) to remain constant over time. There might be other potential confounders such as host genetic and immune response factors, but these data were not available. Moreover, we assumed that the relation between viremia level and platelet count was one-way: viremia level affects platelet count. Whether platelet count affects viremia is unknown.

A notable strength of our study is the utilization of a large pooled dataset, which allowed for a flexible modeling approach to investigate the influence of age, sex, serotype, and immune status, as well as their interactions and nonlinear trends. The use of a model that accounts for left-censored data was needed for capturing the distribution of viremia levels, considering the significant proportion of undetectable values observed.

The study has several limitations. Firstly, the study only included patients from Vietnam, which may restrict the generalisability of the findings to other countries and regions. Secondly, our study assessed viral RNA in the plasma, including both viable and non-viable viral particles, which could potentially lead to an overestimation of the infectious viral particles in the blood. Lastly, data during the incubation period were unavailable as patients are typically not diagnosed with dengue until symptom onset. Therefore, the trajectory of viremia from the time of infection onwards could not be demonstrated, and symptom onset was used as the reference point instead of infection. This could potentially lead to an inaccurate interpretation of the results if the incubation period is influenced by the same factors that we included in our analyses. For example, in cases of secondary infection, the immune response is stronger and quicker than in primary infection. This might lead to a shorter duration between infection and symptom onset, as well as faster viral clearance.

In conclusion, our findings reveal that viremia levels exhibit a rapid decline shortly after symptom onset, becoming undetectable after approximately one week in the majority of patients. Viremia kinetics display variations depending on the infecting serotype. Higher viremia levels are associated with a subsequent reduction in platelet count on illness days 6-8, and an increased risk of experiencing more severe clinical outcomes. Viremia serves as an important predictor of dengue outcomes, independent of the host immune status. However, when considering our previous analysis that involved single early viremia measurements, the addition of daily viremia measurements may not enhance the prediction of worse clinical outcomes. Thus, the measurement of viremia levels during the early febrile phase is an important marker for clinical practice and dengue-related research. Moreover, a faster decline in viremia was found to be associated with a reduced risk of more severe clinical outcomes, suggesting that viremia kinetics could be a good surrogate endpoint for phase-2 dengue therapeutic trials.

## Data Availability

The data that support the findings of this study are available from the corresponding author, (Nguyen Lam Vuong), upon reasonable request.

## Acknowledgements

We thank the staff from the Dengue group at the Oxford University Clinical Research Unit, Hospital for Tropical Diseases at Ho Chi Minh City (Vietnam) and the many hospitals and clinics who recruited and followed patients in the three studies. We gratefully acknowledge the patients and their relatives for participating in these studies.

Additional information

## Funding

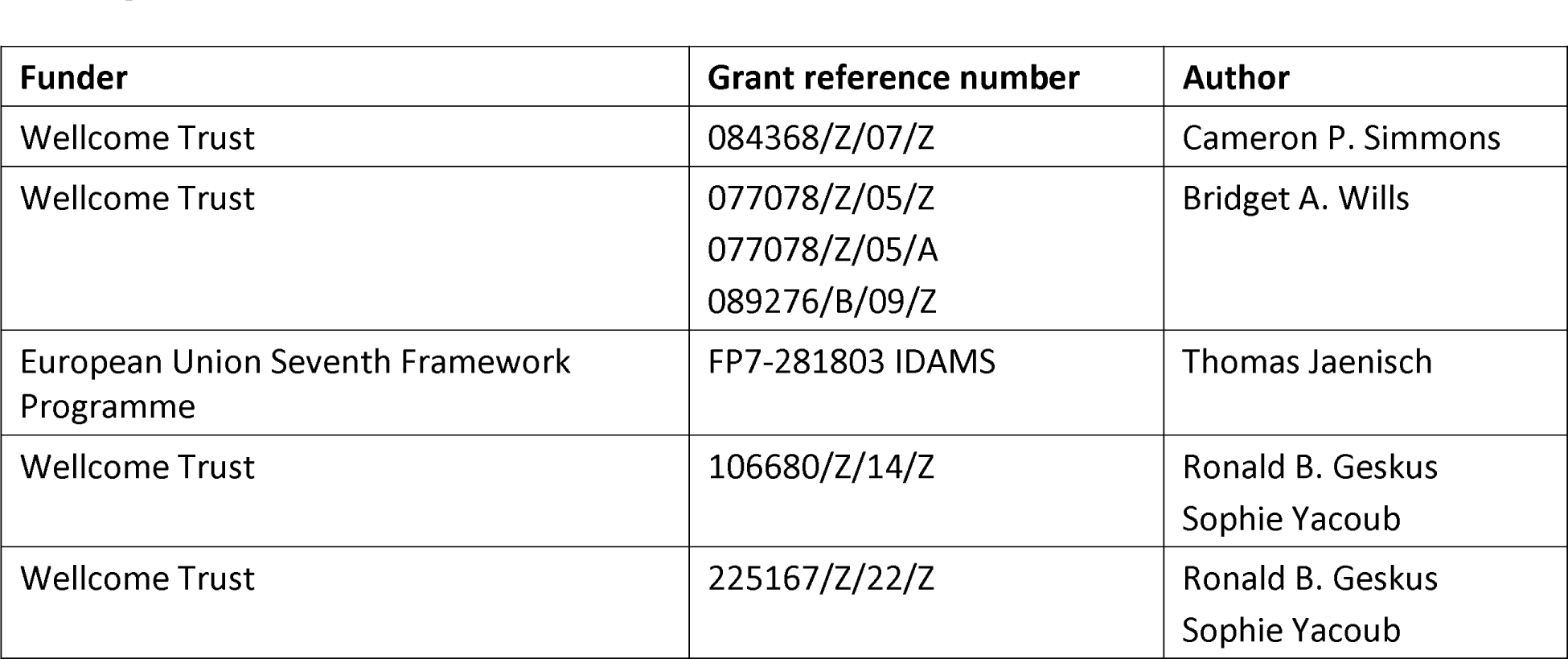

The funders had no role in study design, data collection and interpretation, or the decision to submit the work for publication. For the purpose of open access, the author has applied a CC BY public copyright licence to any Author Accepted Manuscript version arising from this submission.

## Author contributions

Nguyen Lam Vuong: Conceptualization, Methodology, Software, Formal analysis, Data curation, Writing-original draft, Writing-review & editing, Viasualization

Nguyen Than Ha Quyen: Investigation, Resources, Writing-review & editing

Nguyen Thi Hanh Tien: Investigation, Resources, Writing-review & editing

Duong Thi Hue Kien: Investigation, Resources, Writing-review & editing

Huynh Thi Le Duyen: Investigation, Resources, Writing-review & editing

Phung Khanh Lam: Methodology, Investigation, Resources, Data curation, Writing-review & editing, Supervision

Dong Thi Hoai Tam: Conceptualization, Investigation, Resources, Writing-review & editing

Tran Van Ngoc: Conceptualization, Investigation, Resources, Writing-review & editing

Thomas Jaenisch: Conceptualization, Methodology, Investigation, Writing-review & editing, Funding acquisition

Cameron P. Simmons: Conceptualization, Methodology, Investigation, Writing-review & editing, Funding acquisition

Sophie Yacoub: Conceptualization, Methodology, Investigation, Writing-review & editing, Supervision, Funding acquisition

Bridget A. Wills: Conceptualization, Methodology, Investigation, Writing-review & editing, Supervision, Funding acquisition

Ronald B. Geskus: Conceptualization, Methodology, Investigation, Writing-review & editing, Supervision, Funding acquisition

## Ethics

Human subjects: The studies and the blood sample analysis were approved by the Scientific and Ethics Committees of all study sites and by the Oxford Tropical Research Ethics Committee (OxTREC Ref No 017-02; 012-05; and 502-18).

