## Supplementary file for "Dengue Viremia Kinetics and Effects on Platelet Count and Clinical Outcomes: An Analysis of 2340 Patients from Vietnam"

**Appendix 1. Protocol synopses for the three studies**

The three studies described below were performed as part of the longstanding collaboration between the Oxford University Clinical Research Unit and the Hospital for Tropical Diseases in Ho Chi Minh City, Vietnam. All studies were approved by the Scientific and Ethics Committee of the hospital and by the Oxford Tropical Research Ethics Committee.

**Study A**

Title: A study of early dengue disease among children in the community

Study objectives: this study was part of a larger programme of work aiming to a) estimate the burden of paediatric dengue disease in HCMC, b) explore the mechanisms of the endothelial dysfunction associated with dengue by detailed study of well characterized patients throughout the evolution of the disease and c) investigate whether commonly available laboratory parameters (haematocrit, platelets, urine protein) measured during the febrile period can be used to predict subsequent disease severity.

Study period: 2000-2009

Study subjects: previously healthy children aged 5-15 years, presenting to hospital outpatient clinics or to one of several local health centers in Ho Chi Minh City on day 1 or 2 of a non-specific febrile illness. Children were followed daily in the community. In the event of admission to hospital, patients continued to be followed either on the dengue ward or on PICU of the Hospital for Tropical Diseases in HCMC, according to clinical severity.

Study setting: parents/guardians of eligible patients were informed about the study and gave consent. After enrollment baseline blood and urine samples were obtained. Patients were then asked to return each day for 7 days or until full recovery, and then at 1-2 weeks and 1-2 months for follow up. The study nurse assessed each child in the clinic every day, obtained daily blood and urine samples, and requested a medical opinion if there were any concerns about the child or the daily blood results. Two trained study doctors supervised the nurses in the clinics and followed any patients admitted to the hospital. Management in the clinic or referral for hospital admission was at the discretion of the clinic doctors, in consultation with the dengue study doctors.

Data collection: demographic and clinical data were collected on each patient at study entry using a standard case report form. Clinical progress was documented each day specifically focusing on the occurrence of bleeding manifestations or signs of vascular leakage. There was no systematic data collection regarding organ dysfunction as the clinicians felt that these problems were rare in this age group, and no biochemistry (eg liver/renal function) test results were documented in the research files.

Laboratory confirmation of dengue: either positive reverse transcription polymerase chain reaction (RT-PCR) or seroconversion by ELISA (IgM or IgG or both). Testing for dengue non-structural protein 1 (NS1) was performed occasionally but was not part of the original study protocol.

Publications:

1. Dung NT, Duyen HT, Thuy NT, et al. Timing of CD8+ T cell responses in relation to commencement of capillary leakage in children with dengue. *J Immunol* 2010; **184**(12):7281-7287
2. Duyen HT, Ngoc TV, Ha do T, et al. Kinetics of plasma viremia and soluble nonstructural protein 1 concentrations in dengue: differential effects according to serotype and immune status. *J Infect Dis* 2011; **203**(9):1292-1300
3. Hanh Tien NT, Lam PK, Duyen HT, et al. Assessment of microalbuminuria for early diagnosis and risk prediction in dengue infections. *PLoS One* 2013; **8**(1):e54538

**Study B**

Title: Mild Dengue Study

Study objectives: this study was part of a larger programme of work aiming to a) estimate the burden of paediatric dengue disease in HCMC, b) explore the mechanisms of the endothelial dysfunction associated with dengue by detailed study of well characterized patients throughout the evolution of the disease and c) investigate whether commonly available laboratory parameters measured during the febrile period can be used to predict subsequent disease severity.

Study period: 2000-2009

Study subjects: children 5-15 years presenting to the Hospital for Tropical Diseases in HCMC with a febrile illness consistent with possible dengue, who required hospital admission to the paediatric dengue ward, NOT those admitted to HDU/ICU. Only children admitted from home were included (i.e. not transfers from other hospitals).

Study setting: parents/guardians of eligible patients were informed about the study and gave consent. All patients were followed daily until discharge with simple study notes and a daily full blood count (plus any other tests clinically indicated), and treated according to individual diagnoses. On the day of study enrolment all patients had a plasma sample obtained for research investigations. On the 6^th^ illness day a second plasma sample was obtained. At discharge the notes were reviewed and each child was assigned a final diagnosis and clinical disease category, using carefully defined severity criteria for vascular leak and bleeding. The patients were asked to return for follow up at one month, and again at two months if there were any ongoing concerns.

Data collection: demographic and clinical data were collected on each patient at study entry using a standard case report form. Clinical progress was documented each day specifically focusing on the occurrence of bleeding manifestations or signs of vascular leakage. There was no systematic data collection regarding organ dysfunction as the clinicians felt that these problems were rare in this age group, and no biochemistry (eg liver/renal function) test results were documented in the research files.

Laboratory confirmation of dengue: either positive RT-PCR or seroconversion by ELISA (IgM or IgG or both). NS1 testing was not performed.

Publications:

1. Dung NT, Duyen HT, Thuy NT, et al. Timing of CD8+ T cell responses in relation to commencement of capillary leakage in children with dengue. *J Immunol* 2010; **184**(12):7281-7287
2. Trung DT, Thao LTT, Dung NM, et al. Clinical Features of Dengue in a Large Vietnamese Cohort: Intrinsically Lower Platelet Counts and Greater Risk for Bleeding in Adults Than Children. *PLoS Negl Trop Dis* 2012; **6**(6):e1679.
3. Lam PK, Ngoc TV, Thuy TT, et al*.* The value of daily platelet counts for predicting dengue shock syndrome: Results from a prospective observational study of 2301 Vietnamese children with dengue. *PLoS Negl Trop Dis* 2017; **11**(4): e0005498
4. Hoang Quoc C, Henrik S, Isabel RB, et al. Synchrony of Dengue Incidence in Ho Chi Minh City and Bangkok. *PLoS Negl Trop Dis* 2016; **10**(12):e0005188

**Study C**

Title: IDAMS – observational study in early dengue (ClinicalTrials.gov Identifier: NCT01550016)

Study objectives: to improve diagnosis and clinical management of dengue through approaches designed a) to differentiate between dengue and other common febrile illness within 72 h of fever onset, and b) among patients with dengue to identify markers predictive of the likelihood of evolving to a more severe disease course.

Study period: 2011-2016

Study subjects: both adults and children (≥5 years) were eligible for enrolment. This was a prospective multi-centre observational study that enrolled approximately 7,500 patients presenting with a febrile illness consistent with possible dengue to outpatient health facilities in urban centres in eight countries across Asia and Latin America. Following appropriate informed consent, subjects presenting at one of the designated sites with fever for ≤ 72 hours without localizing features, i.e. consistent with a possible diagnosis of dengue, were enrolled.

Study setting: Following enrolment, clinical history and examination findings were recorded in the case report form and a 3–5 ml (age-dependent) research blood sample was obtained, together with appropriate samples to measure a range of haematological and biochemical parameters in line with local laboratory capacity. Patients were then reviewed daily in the OPD until fully recovered and afebrile for 24 hours, or for up to six days from enrolment. A full blood count was carried out each day, and on the last acute illness visit (within approximately 24 hours of defervescence) a second sample for a biochemical profile was obtained together with a sample for serology. All patients were then asked to attend a final follow-up visit around day 10-14 of illness, at least one week from the last visit during the acute illness. All management decisions throughout the acute illness were at the discretion of the clinic physicians. Any patient requiring hospital admission continued to be followed daily using a similar but more detailed CRF, with the indication(s) for admission documented, and all management interventions recorded together with the physician’s rationale for these interventions.

Data collection: a structured clinical questionnaire was completed upon enrolment and then once daily for up to six days for all patients in the study. This CRF included detailed clinical signs and symptoms, as well as all standard laboratory results including liver and renal function.

Laboratory confirmation of dengue: any case with virological evidence of dengue as shown by a positive RT-PCR assay or NS1 ELISA test, was defined as having laboratory-confirmed dengue.

Publications:

1. Nguyet MN, Duong TH, Trung VT, et al. Host and viral features of human dengue cases shape the population of infected and infectious Aedes aegypti mosquitoes. *Proc Natl Acad Sci USA* 2013; **110**(22):9072-9077
2. Jaenisch T, Tam DTH, Kieu NTT, et al. Clinical evaluation of dengue and identification of risk factors for severe disease: protocol for a multicentre study in 8 countries. *BMC Infect Dis* 2016; **16**: 120
3. Vuong NL, Le Duyen HT, Lam PK, et al. C-reactive protein as a potential biomarker for disease progression in dengue: a multi-country observational study. *BMC Med* 2020; 18(1): 35
4. Vuong NL, Lam PK, Ming DKY, et al. Combination of inflammatory and vascular markers in the febrile phase of dengue is associated with more severe outcomes. *eLife* 2021; 10: e67460

**Appendix 2. Definitions used for dengue diagnostics and clinical severity classification**

**Dengue diagnostics**

Criteria for dengue diagnostics in the three studies were harmonized. Laboratory-confirmed dengue was defined by either a positive reverse transcription polymerase chain reaction (RT-PCR) or a positive dengue NS1 antigen test on plasma samples.

**Immune status**

A probable primary infection was defined by two negative/equivocal IgG results on two samples taken at least two days apart during the first ten days after symptom onset, with at least one sample obtained during the convalescent phase (illness days 6-10).

A probable secondary infection was defined by at least one positive IgG result during the first ten days.

In cases without time-appropriate IgG results, immune status was classified as indeterminate.

**Clinical severity classification**

| **Outcome** | **Definition** |
| --- | --- |
| Severe dengue | One or more of the following:   1. Severe plasma leakage resulting in dengue shock syndrome and/or respiratory distress due to fluid accumulation 2. Severe bleeding 3. Severe organ impairment |
|  | Dengue Shock Syndrome: Pulse pressure (difference between systolic and diastolic pressures) ≤ 20 mmHg or hypotension for age plus signs of poor capillary perfusion (cold extremities, delayed capillary refill, or rapid pulse rate). Clinician’s assessment was accepted for occasional cases where the PP was between 20-25. |
|  | Respiratory Distress: Increased respiratory rate for age, with signs of increased work of breathing (retractions, nasal flaring, accessory muscle use) and need for additional support such as oxygen supplementation, CPAP or intubation |
| Plasma leakage | 1. Haemoconcentration. Defined as ≥ 20% increase in hematocrit (HCT) from baseline (minimum HCT within illness days 1-3) to acute phase (maximum HCT within illness days 4-7) 2. And/or evidence of fluid accumulation (pleural/peritoneal) on ultrasound or x-ray |
|  | Severe: Dengue shock syndrome and/or respiratory distress due to plasma leakage |
|  | Moderate: Evidence of plasma leakage but never developed shock or respiratory distress |
|  | None: Haemoconcentration < 20% (definition as above) and no evidence of fluid accumulation if chest X-ray/ultrasound done |
|  | Indeterminate: Missing data for HCT (either baseline or acute phase or both) |
| Severe bleeding | One or more of the following:   1. Any bleeding leading to hemodynamic instability 2. Any bleeding resulting in death or permanent disability 3. Any bleeding into a critical organ (e.g., central nervous system bleed) 4. Any bleeding that results in need for blood transfusion 5. Any bleeding that persists after measures are taken to stop bleeding (e.g., application of pressure) AND patient requires more intensive monitoring in an ICU or HDU |
| Severe organ impairment | Assessments were not systematic. For patients investigated on the basis of the attending clinician’s concerns the following definitions were applied. |
|  | Severe liver involvement: An acute clinical syndrome consistent with acute hepatitis, with new onset jaundice  or coagulopathy (INR≥1.5) or encephalopathy. |
|  | Severe neurological involvement: Any new onset acute neurological signs or symptoms, except occurrence of a single simple febrile convulsion with full recovery within 30 minutes. |
|  | Severe renal involvement: Increase in creatinine to more than 1.5 times the upper limit of normal for age, without existing kidney disease. |

Note that the criteria used for classifying dengue severity are derived from the World Health Organization guidelines [1] and standard endpoint definitions for dengue trials [2]. These criteria have been adjusted to align with the actual circumstances and the data accessible for the studies included in this analysis.

**Appendix 3. Statistical analysis**

We constructed a directed acyclic graph (DAG) to outline the hypothesized pathways from viremia level to platelet count and clinical outcomes.

**Appendix 3-figure 1. Directed Acyclic Graph illustrating the presumed causal relationships among variables**

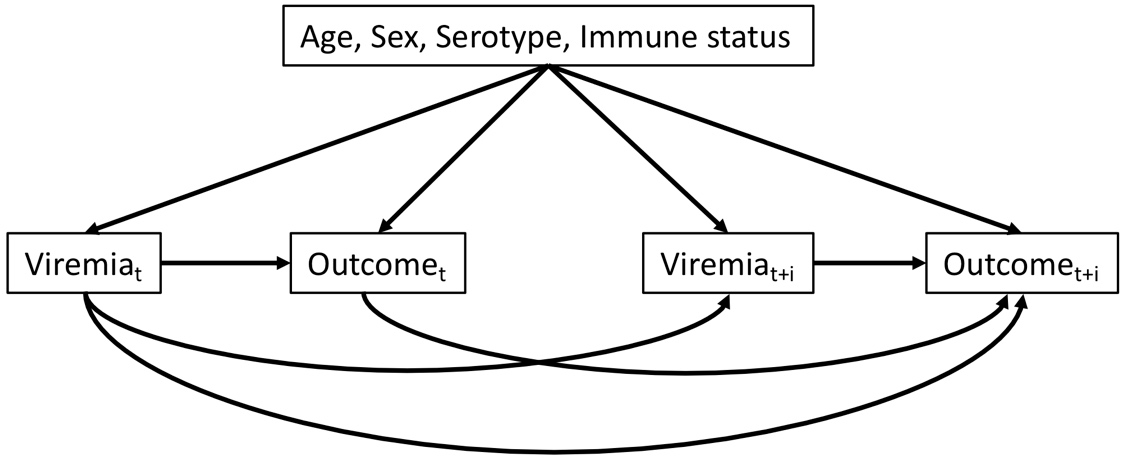

*t: illness day t (from day 1 to 7)*

*t+i: after day t (i ≥ 1)*

*Outcome can be platelet count or clinical outcomes (severe dengue or plasma leakage)*

**Analysis #1: Viremia kinetics and association with clinical characteristics**

In the one-step PCR cohort, viremia levels were left-censored at specific detection limits: 300 copies/ml for DENV-1 and DENV-3, 60 copies/ml for DENV-2, and 600 copies/ml for DENV-4.

For the two-step PCR cohort, viremia levels were left-censored at 1000 copies/ml for DENV-1, DENV-2, and DENV-3. Due to the limited number of cases (two patients), DENV-4 was excluded from this cohort. We chose 1000 copies/ml as the detection limit for the two-step PCR method (the old method) based on assumptions of its higher detection limit compared to the one-step PCR method (the new method) which has been validated.

To account for non-linear trends, splines were used for age (with three knots at 7, 13, and 28 years) and illness day (with four knots at day 1, 2, 4, and 6).

To fit the linear mixed-effects models with censored outcome, a Bayesian framework using Markov chain Monte Carlo (MCMC) methods was implemented using the R package 'MCMCglmm'. The models were run for a total of 20,000 MCMC iterations. Model adequacy was assessed through parameter trace plots and empirical distribution estimation [1] from the models, indicating satisfactory fit for both models for log-10 viremia and 10th-root viremia.

**Appendix 3-figure 2. Empirical distribution estimation obtained from viremia kinetics model**
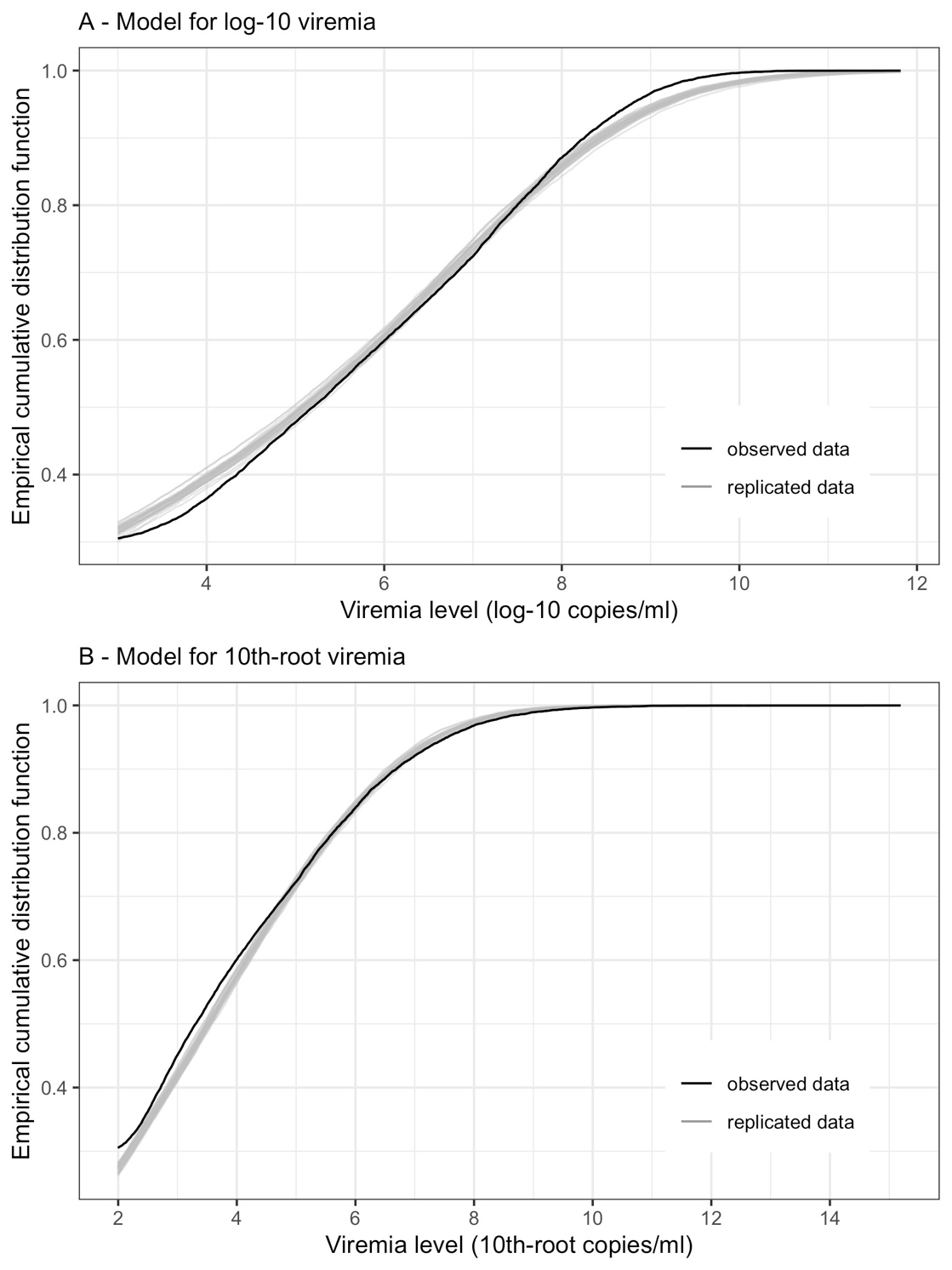

*The figures show the empirical cumulative distribution function of the observed viremia levels (represented by the black curve), along with the empirical cumulative distribution function derived from 100 simulated sets of viremia levels generated by the model (represented by the grey curves). The plots are started from 1000 copies/ml (on the original scale). The close resemblance between the observed and simulated distributions in both models based on log-10 and 10th-root transformations of viremia levels suggests that the models provide a relatively good fit to the data.*

The model using 10th-root viremia showed a relatively better fit, particularly for high viremia levels. However, considering the common use of the log transformation of viremia in studies, and the similarity of the predictions between the two models, we decided to use log-10 viremia as the main analysis of this study.

**Analysis #2: Effect of viremia on platelet count**

*Analysis #2a: Compare the effect of viremia level on platelet count on illness days 6 to 8 using linear mixed effects models. Separate models were fitted for viremia level measured 0 to 5 days earlier than the platelet count*

We investigated the effect on platelet count of viremia 0 to 5 days earlier. Since almost all viremia values were undetectable after day 8, data for this analysis was restricted to days 6 to 8 and included patients who had available data on viremia on the same day and viremia 1 to 5 days earlier. We used a linear mixed-effects model for platelet count, including as covariates log-10 viremia, age, sex, serotype, immune status, PCR method, and illness day. We included interactions between illness day and all other covariates, interactions between log-10 viremia level and serotype and immune status, and interaction between serotype and immune status. Potential nonlinear effects of log-10 viremia and age were investigated using splines with 3 knots at the 10^th^, 50^th^, and 90^th^ percentiles. To assess model fit, we examined the log-likelihood, AIC, and BIC values for each model.

**Appendix 3-table 1. Comparison of the models in analysis #2a**

| **Model** | **Degrees of freedom** | **Log-likelihood** | **AIC** | **BIC** |
| --- | --- | --- | --- | --- |
| Viremia on the same day | 55 | -498.5 | 1106.9 | 1377.2 |
| Viremia 1 day earlier | 55 | -486.2 | 1082.3 | 1352.6 |
| Viremia 2 days earlier | 55 | -479.4 | 1068.9 | 1339.1 |
| Viremia 3 days earlier | 55 | -452.5 | 1015.1 | 1285.3 |
| Viremia 4 days earlier | 55 | -469.5 | 1048.9 | 1319.2 |
| Viremia 5 days earlier* | 54 | -478.4 | 1064.8 | 1330.1 |

**The model for viremia 5 days earlier had fewer degrees of freedom due to the absence of a binary variable indicating detectable or undetectable viremia values (as all patients had detectable viremia values 5 days earlier).*

Among the models considered, the model for viremia 3 days earlier demonstrated the best fit.

*Analysis #2b: Compare the effect of viremia level on platelet count using linear regression. Separate models were fitted for platelet count on illness day 2 to 8 and for viremia level measured 0 to 5 days earlier than the platelet count.*

To validate the findings from Analysis #2a, we conducted standard linear regression models to assess the relation between viremia level and platelet count for each combination of platelet count on illness day 2 to 8 and viremia level on day 0 to 5 before the platelet measurement. The models included log-10 viremia (with splines using 3 knots at the 10^th^, 50^th^, and 90^th^ percentiles), the binary variable indicating detectable or undetectable viremia (if applicable), age (with splines using 3 knots at 7, 13, and 28 years), sex, serotype, immune status, and PCR method as covariates. No interaction terms were included due to limited data availability for each illness day. To evaluate model fit, we assessed the log-likelihood value for each model.

**Appendix 3-table 2. Comparison of the models in analysis #2b**

| **Model** | **Day 2** | **Day 3** | **Day 4** | **Day 5** | **Day 6** | **Day 7** | **Day 8** |
| --- | --- | --- | --- | --- | --- | --- | --- |
| Same day | 2.0 (24) | -58.7 (145) | -95.9 (220) | -99.7 (227) | -84.9 (198) | -252.6 (533) | -210.5 (449) |
| 1 day earlier | 0.9 (24) | **-53.1 (134)** | -95.2 (218) | -98.0 (224) | -80.1 (188) | -243.4 (515) | -208.7 (443) |
| 2 days earlier |  | -56.0 (138) | **-88.8 (206)** | -94.0 (216) | -80.9 (190) | -232.8 (494) | -205.2 (438) |
| 3 days earlier |  |  | -91.8 (210) | **-89.8 (208)** | **-76.2 (180)** | **-229.3 (487)** | **-189.6 (407)** |
| 4 days earlier |  |  |  | -92.9 (212) | -78.7 (185) | -241.3 (511) | **-189.6 (407)** |
| 5 days earlier |  |  |  |  | -83.0 (192) | -245.4 (517) | -196.5 (419) |

*Statistics are log-likelihood (AIC). The highest log-likelihood values (along with the lowest AIC) per column are highlighted in bold. Note that all analyses per each column use the same sample size.*

Overall, the models for viremia level 3 days earlier performed the best. These models had the highest log-likelihood values on illness days 5 to 8, and the second-highest log-likelihood values on illness day 4, with values closely approaching the highest log-likelihood values. These results align with the analyses conducted in Analysis #2a.

*Analysis #2c: Effect of day 1-7 viremia on platelet count on day 1-10*

The formula of the supermodel was as follows:

- Fixed effect:	PLT ~ (Viremia + LOD) * Serotype * (Immune + PCR) + Age + Sex + DOI + LM +

DOI : [(Viremia + LOD) * Serotype * PCR + Immune + Age + Sex + LM] +

LM : [(Viremia + LOD) * Serotype * PCR]

- Random effect:	PLT ~ (DOI | id : LM)
  - PLT: platelet count
  - Viremia: splines of log-10 viremia with 3 knots at 4, 6, and 8
  - LOD: the binary variable indicating detectable or undetectable viremia
  - Immune: immune status with 3 levels
  - PCR: PCR method (one-step or two-step)
  - Age: splines of age with 3 knots at 7, 13, and 28
  - DOI: splines of illness day with 3 knots at 2, 5, and 8
  - LM: splines of landmark day with 3 knots at 2, 4, and 6, except for in the random effect, landmark day was treated as a factor variable with 7 levels

The model was well-fitted, demonstrated by the empirical distribution estimation plot generated from the model.

**Appendix 3-figure 4. Empirical distribution estimation obtained from platelet count model**
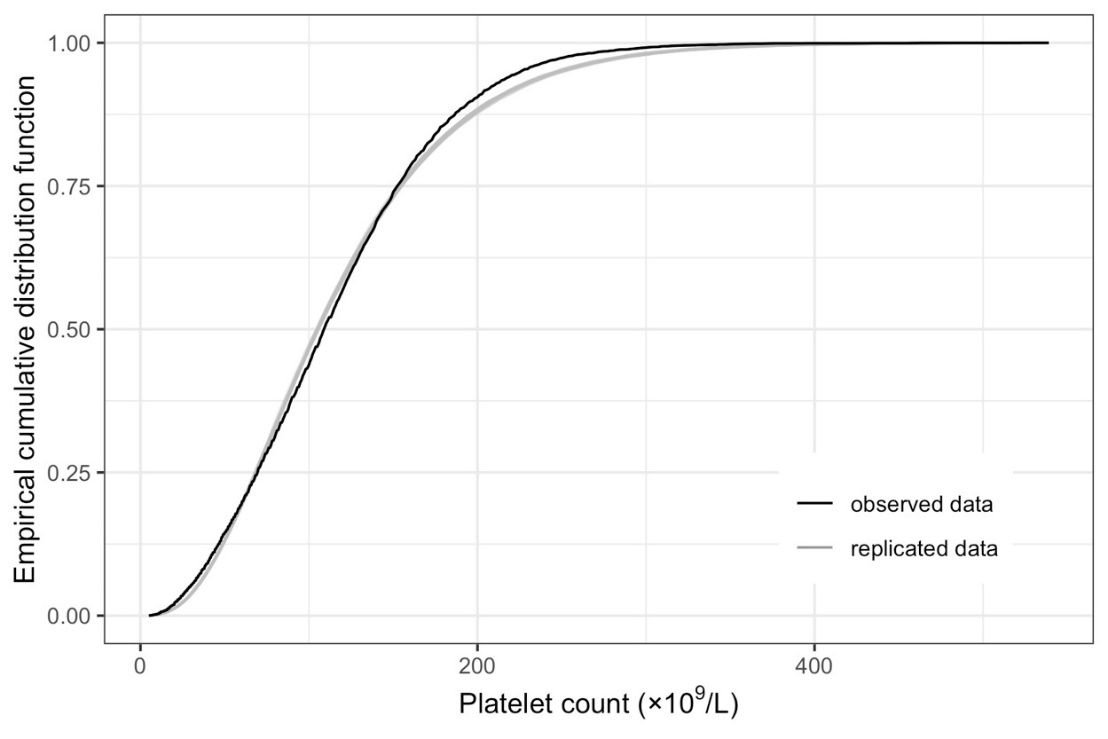

*The figure shows the empirical cumulative distribution function of the observed platelet counts (represented by the black curve), along with the empirical cumulative distribution function derived from 100 simulated data points generated by the model (represented by the grey curves). The close resemblance between the observed and simulated distributions suggests that the model provides a relatively good fit to the data.*

**Analysis #3: Effect of viremia level on clinical outcomes**

The unknown day of plasma leakage for 26 patients was set at day 5.

For the supermodel that combined all landmark data sets, we employed the "independence" covariance structure for the generalized estimating equations (GEE). The covariates included log-10 viremia level, the binary variable describing detectable or undetectable viremia, age, sex, serotype, immune status, study, and landmark illness day. Splines were used to allow for nonlinear trends by log-10 viremia, age, and landmark day. We included the interactions between viremia with all other covariates (age, sex, and study) and the four-way interaction between log-10 viremia with serotype, immune status and landmark day.

Due to the limited number of severe dengue cases (65 in total), the interactions allowed in the models for severe dengue and plasma leakage differed:

- For severe dengue, we did not include any interactions between the binary variable describing detectable viremia with other covariates. Log-10 viremia and landmark day were specified as a linear trend in the interactions.
- For plasma leakage, we additionally included the interactions between the binary variable describing detectable viremia with all other covariates (age [as a linear trend], sex, serotype, immune status, study, and landmark illness day [as a linear trend]). In the interactions between log-10 viremia and others, log-10 viremia was specified as a linear trend, whereas age and landmark day was specified with splines (similarly to in the main terms).

The formula of the supermodels was as follows:

- Severe dengue ~ Viremia_[splines]_ + LOD + LM_[splines]_ + Serotype * Immune + Age_[splines]_ + Sex + Study +

Viremia_[linear]_ : (LM_[splines]_ * Serotype * Immune + Age_[splines]_ + Sex + Study)

- Plasma leakage ~ Viremia_[splines]_ + LOD + LM_[splines]_ + Serotype * Immune + Age_[splines]_ + Sex + Study +

Viremia_[linear]_ : (LM_[splines]_ * Serotype * Immune + Age_[splines]_ + Sex + Study) +

LOD : (LM_[linear]_ + Serotype + Immune + Age_[linear]_ + Sex + Study)

- Viremia_[splines]_: splines of log-10 viremia with 3 knots at 4, 6.4, and 8.5
- Viremia_[linear]_: linear trend of log-10 viremia
- LOD: the binary variable indicating detectable or undetectable viremia
- Immune: immune status
- Age_[splines]_: splines of age with 3 knots at 7, 13, and 28
- Age_[linear]_: linear trend of age
- LM_[splines]_: splines of landmark day with 3 knots at 2, 4, and 6
- LM_[linear]_: linear trend of landmark day

Some missing data were present for plasma leakage and immune status (i.e., the indeterminate immune status). In the complete-case analysis, individuals with missing plasma leakage were excluded, and immune status was analyzed with three groups, including indeterminate status. In the imputed data analysis, we used results from the imputation performed in the previous analysis of the effect of viremia in the early febrile phase on clinical outcomes [2]. Logistic regression was the imputation model for immune status and plasma leakage. All available baseline data were included in the imputation model, including study, age, gender, weight, enrolment date, illness day at enrolment, serotype, immune status, haematocrit and platelet count at enrolment, and the outcomes (hospitalization along with its characteristics [illness day when hospitalized and length of hospital stay], plasma leakage, and severe dengue). Twenty imputed datasets were generated, and 25 cycles per dataset were performed. Rubin’s rules were applied to pool estimates from the logistic regression models over the imputed datasets.

### Appendix 4. Descriptive analysis

**Appendix 4-figure 1. Distribution of log-10 and 10th-root viremia levels by illness day**

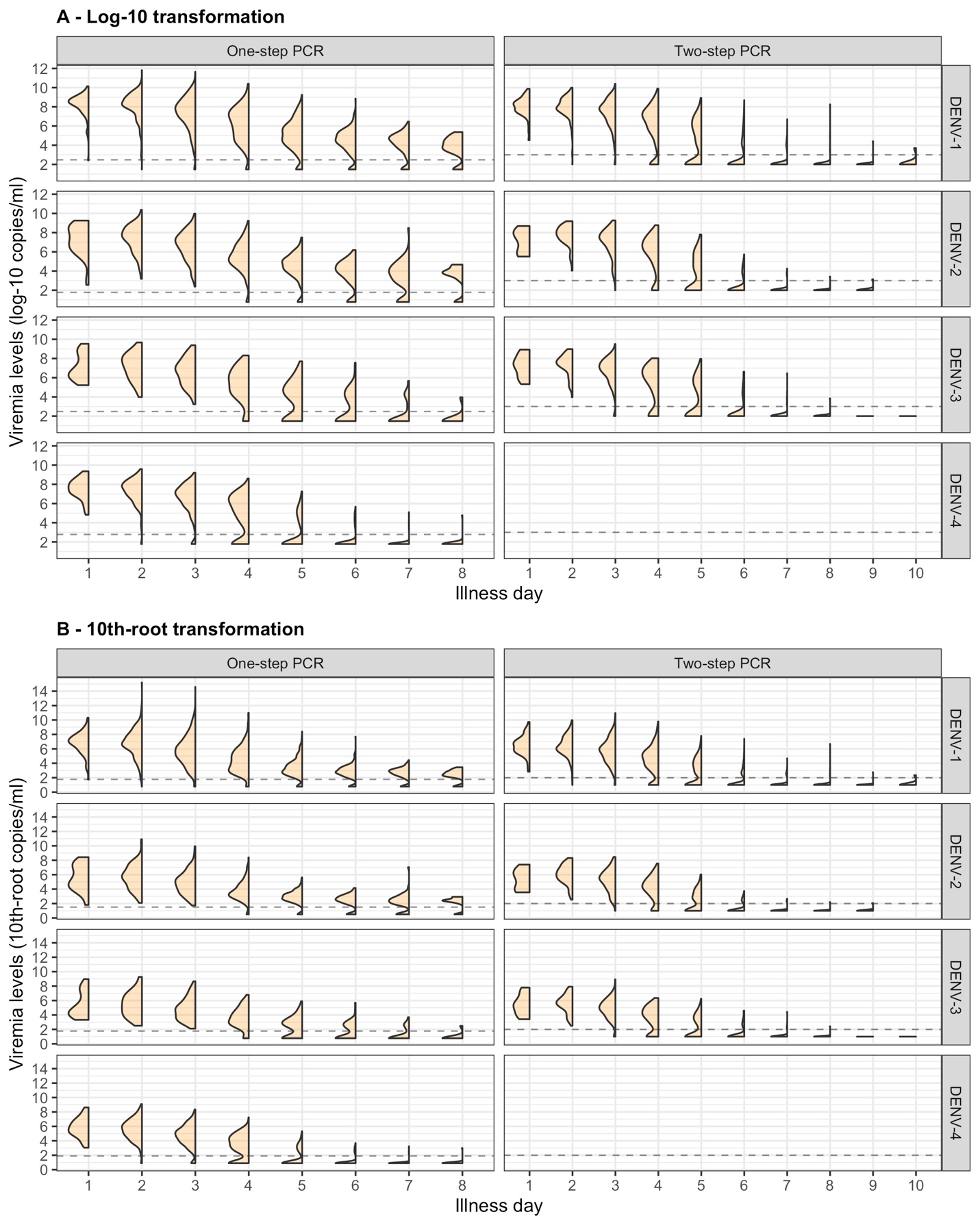

Values below the detection limit are set as 1/2 of the detection limit in each corresponding transformation scale.

### Appendix 4-figure 2. Distribution and individual trajectory of platelet count

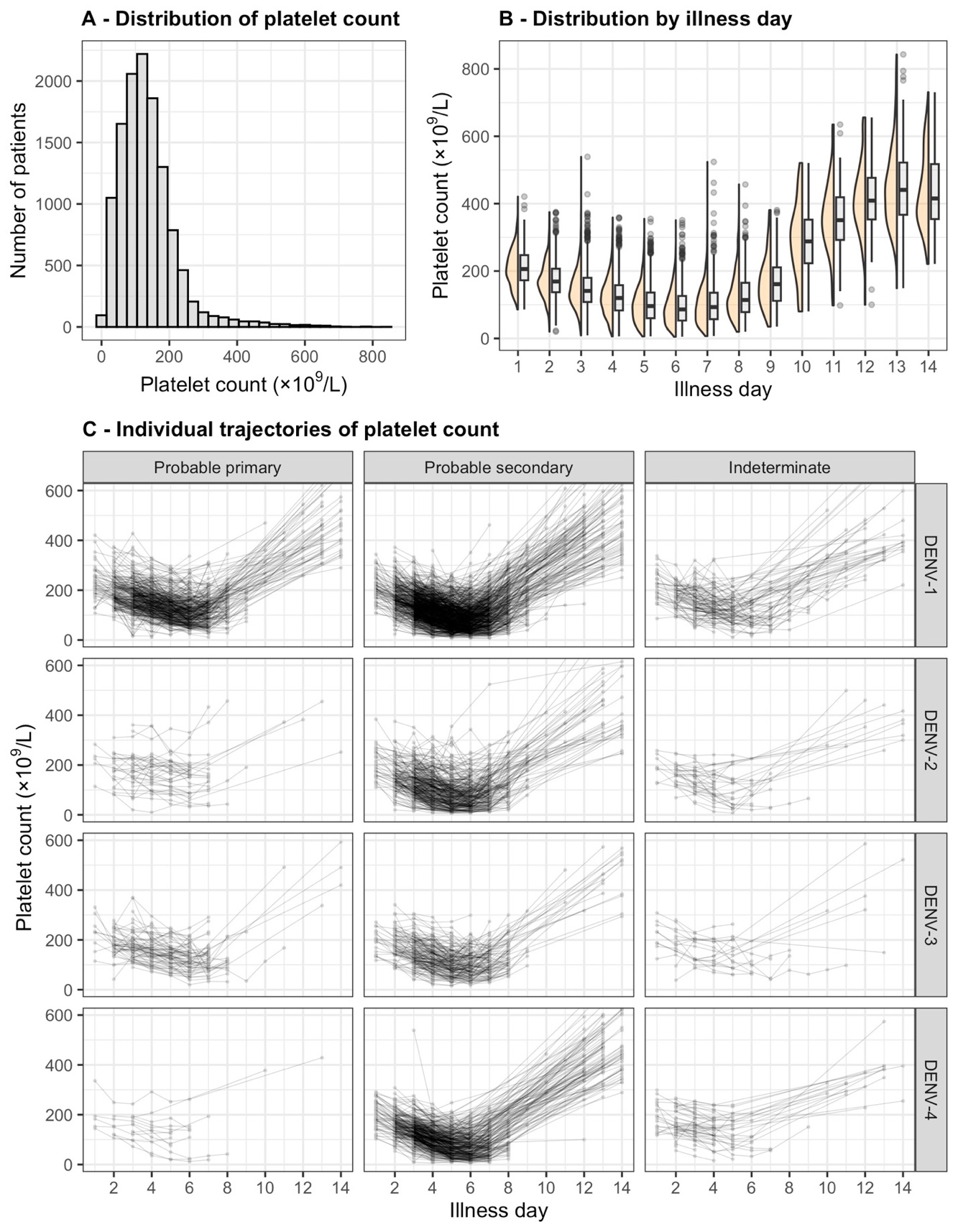

### The distribution of platelet count is shown with all measurements (A) and by illness day (B). In (B), the line inside each box represents the median, the upper and lower margins of each box indicate the interquartile range (25th; 75th percentile), and the violin curves illustrate the distribution of platelet count. Individual trajectories of platelet count are shown in (C): the dots represent the measured platelet counts and are connected by lines for each individual patient. The summary is restricted up to illness day 14. DENV, dengue virus.

**Appendix 4-table 1. Distribution of clinical outcomes by serotype and immune status**

|  |  | **Severe dengue^#^** | |  | **Plasma leakage^#^** | | |
| --- | --- | --- | --- | --- | --- | --- | --- |
|  | **N^$^** | **No**  **(n=2275)** | **Yes**  **(n=65)** |  | **No**  **(n=1935)** | **Yes**  **(n=353)** | **Indeterminate**  **(n=52)** |
| **DENV-1** | **1264** | **1224 (96.8)** | **40 (3.2)** |  | **1028 (81.3)** | **200 (15.8)** | **36 (2.8)** |
| Probable primary | 356 (28.2) | 355 (99.7) | 1 (0.3) |  | 307 (86.2) | 35 (9.8) | 14 (3.9) |
| Probable secondary | 774 (61.2) | 737 (95.2) | 37 (4.8) |  | 603 (77.9) | 151 (19.5) | 20 (2.6) |
| Indeterminate | 134 (10.6) | 132 (98.5) | 2 (1.5) |  | 118 (88.1) | 14 (10.4) | 2 (1.5) |
| **DENV-2** | **373** | **358 (96.0)** | **15 (4.0)** |  | **292 (78.3)** | **75 (20.1)** | **6 (1.6)** |
| Probable primary | 37 (9.9) | 37 (100.0) | 0 (0.0) |  | 29 (78.4) | 7 (18.9) | 1 (2.7) |
| Probable secondary | 299 (80.2) | 286 (95.7) | 13 (4.3) |  | 232 (77.6) | 63 (21.1) | 4 (1.3) |
| Indeterminate | 37 (9.9) | 35 (94.6) | 2 (5.4) |  | 31 (83.8) | 5 (13.5) | 1 (2.7) |
| **DENV-3** | **252** | **250 (99.2)** | **2 (0.8)** |  | **208 (82.5)** | **34 (13.5)** | **10 (4.0)** |
| Probable primary | 68 (27.0) | 68 (100.0) | 0 (0.0) |  | 58 (85.3) | 8 (11.8) | 2 (2.9) |
| Probable secondary | 165 (65.5) | 163 (98.8) | 2 (1.2) |  | 132 (80.0) | 25 (15.2) | 8 (4.8) |
| Indeterminate | 19 (7.5) | 19 (100.0) | 0 (0.0) |  | 18 (94.7) | 1 (5.3) | 0 (0.0) |
| **DENV-4** | **451** | **443 (98.2)** | **8 (1.8)** |  | **407 (90.2)** | **44 (9.8)** | **0 (0.0)** |
| Probable primary | 13 (2.9) | 13 (100.0) | 0 (0.0) |  | 13 (100.0) | 0 (0.0) | 0 (0.0) |
| Probable secondary | 381 (84.5) | 373 (97.9) | 8 (2.1) |  | 337 (88.5) | 44 (11.5) | 0 (0.0) |
| Indeterminate | 57 (12.6) | 57 (100.0) | 0 (0.0) |  | 57 (100.0) | 0 (0.0) | 0 (0.0) |

Summary statistics are n (%).

^$^The percentages of immune status are computed by the total sample size within each DENV serotype.

^#^The percentages are computed by the total sample size per row.

DENV, dengue virus

**Appendix 4-figure 3. Difference in log-10 viremia from first measurement**

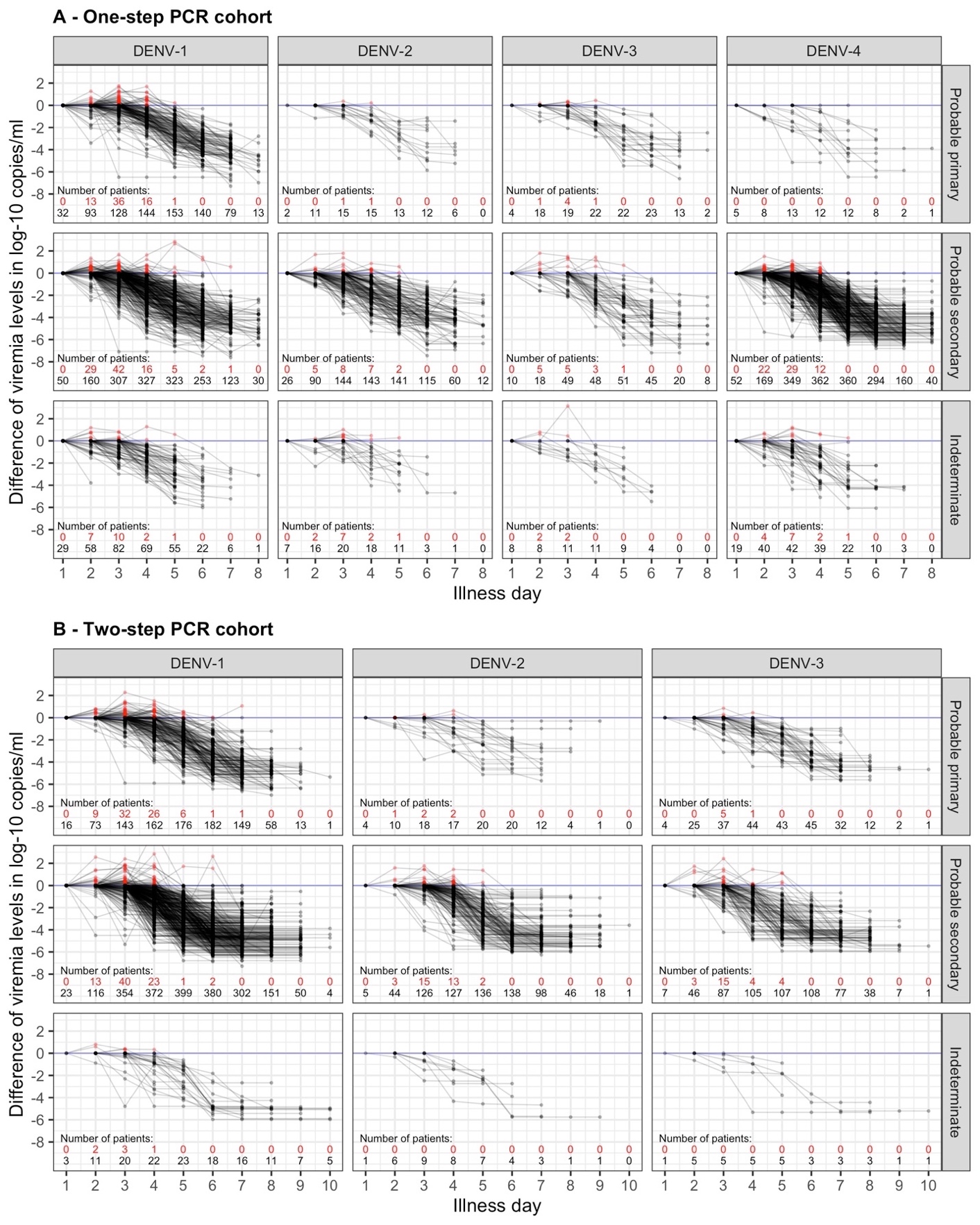

The red dots represent values that are higher than the first value. The red and black numbers indicate the number of measurements that are higher and lower than the first value, respectively. The upper limit of the y-axis is set at 2.5 to provide a clearer visualization. Values below the detection limit are set as the corresponding detection limit to calculate the differences. DENV, dengue virus; PCR, polymerase chain reaction.

### Appendix 5. Results for viremia kinetics and relationship with clinical characteristics

### Appendix 5-figure 1. Compare fitted trends in mean viremia levels between one-step and two-step PCR methods

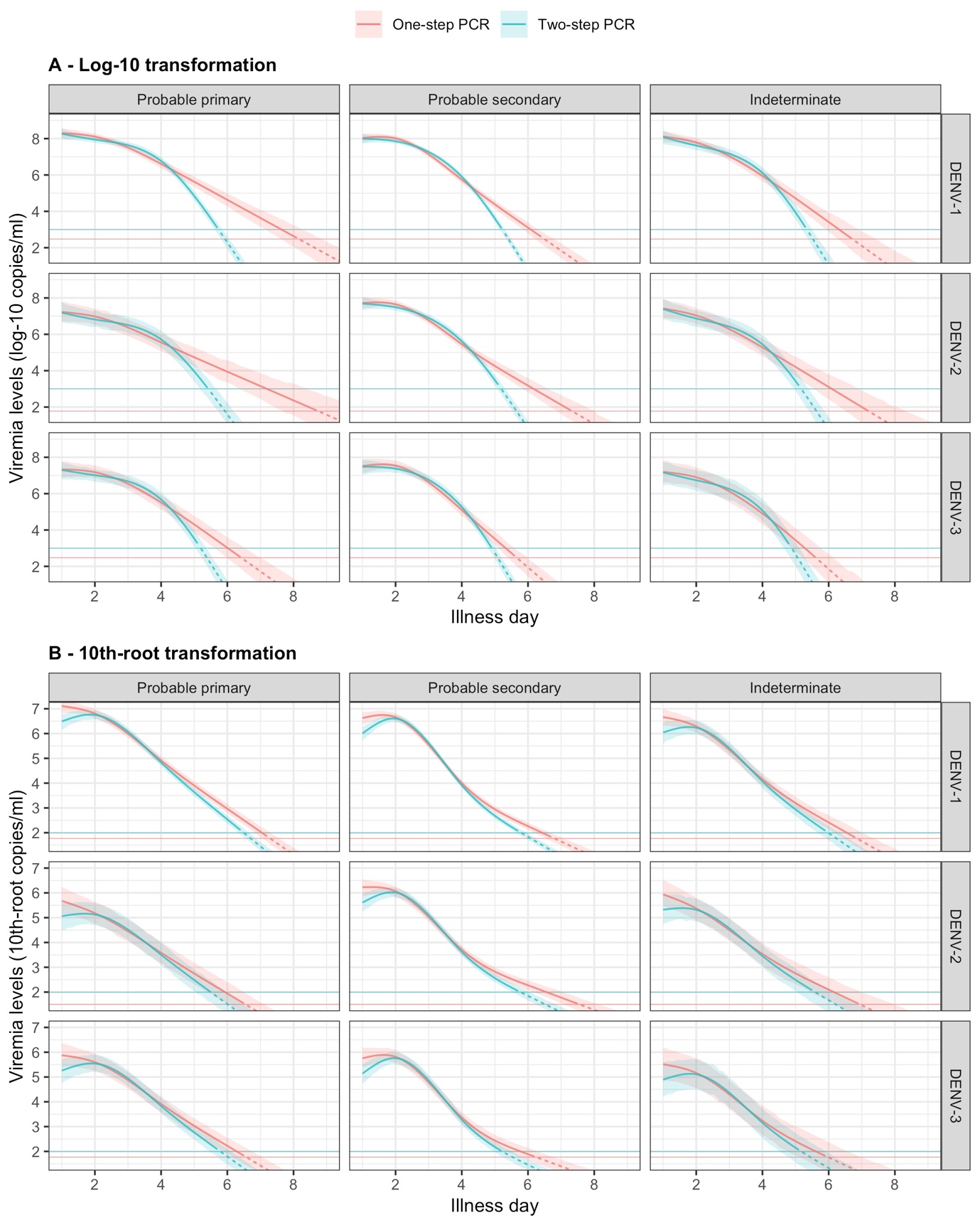

The estimated mean viremia levels are represented by colored lines. The 95% credible intervals are shown as colored shaded regions. The horizontal lines indicate the detection limits. Dashed lines indicate the estimated viremia levels below the detection limits. Viremia levels are shown for age of 10 years and male sex. DENV, dengue virus; PCR, polymerase chain reaction.

### Appendix 5-table 1. P-values of parameters in the models of viremia

| **Parameter** | **P value** |
| --- | --- |
| Age (overall effect) | <0.0001 |
| - All interactions of Age | 0.0015 |
| - Nonlinear effect of Age | <0.0001 |
| Sex (overall effect) | 0.0008 |
| - All interactions of Sex | 0.0011 |
| Serotype (overall effect) | <0.0001 |
| - All interactions of Serotype | <0.0001 |
| Immune status (overall effect) | <0.0001 |
| - All interactions of Immune status | <0.0001 |
| PCR method (overall effect) | <0.0001 |
| - All interactions of PCR method | <0.0001 |
| Day (overall effect) | <0.0001 |
| - All interactions of Day | <0.0001 |
| - Nonlinear effect of Day | <0.0001 |
| Serotype * Immune status | 0.0066 |
| Age * Day | 0.0015 |
| Sex * Day | 0.0011 |
| Serotype * Day | <0.0001 |
| Immune status * Day | <0.0001 |
| PCR * Day | <0.0001 |

P values are calculated using the Wald test from the R package ‘aod’. This test computes the Wald *chi-squared* test for 1 or more coefficients given their variance-covariance matrix from the model.

A * B is the interaction between A and B.

PCR, polymerase chain reaction

### Appendix 5-figure 2. Fitted trends in mean viremia levels by models based on 10th-root and log-10 viremia transformations

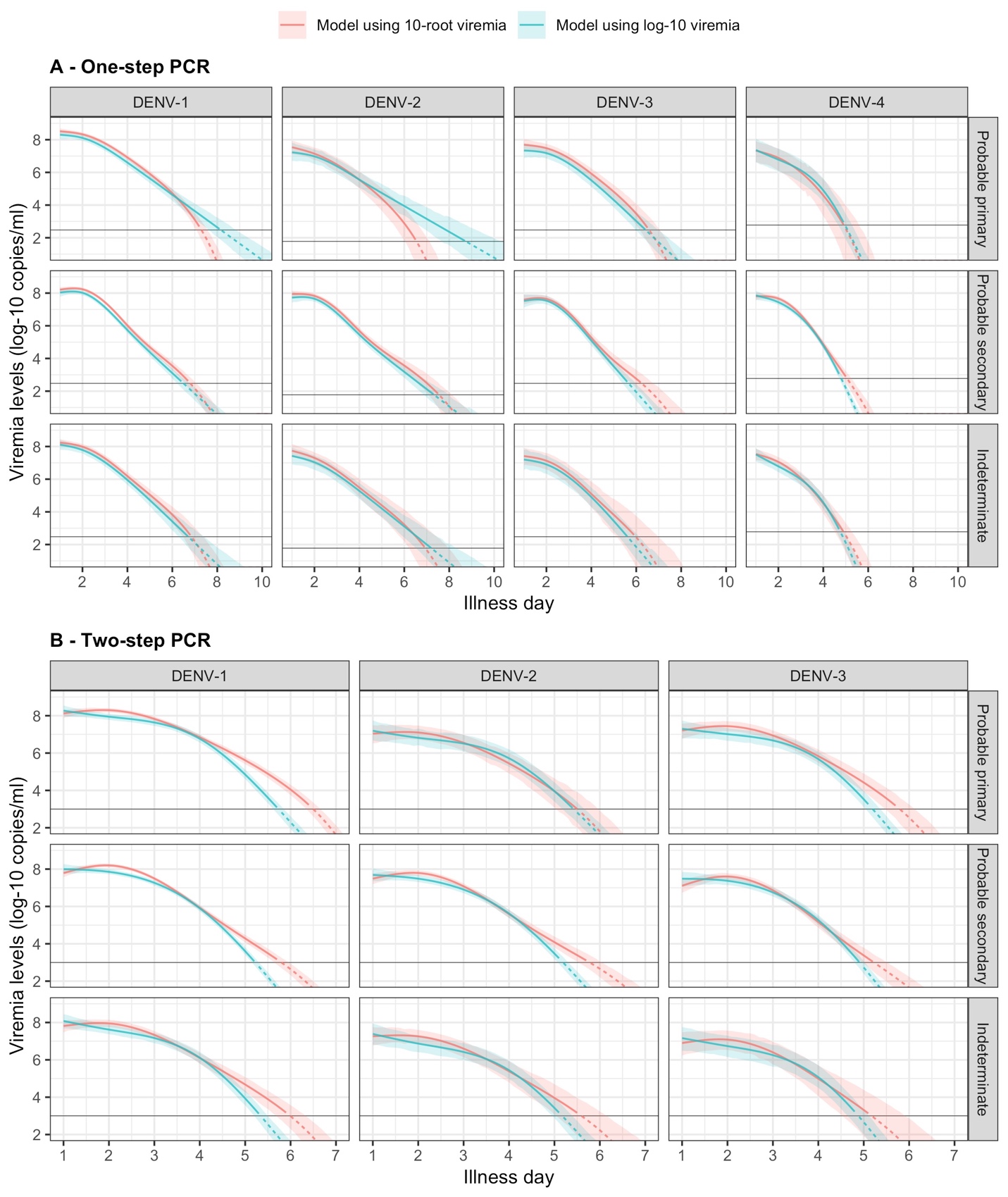

The estimated mean viremia levels are represented by colored lines. The 95% confidence intervals are shown as colored shaded regions. The horizontal lines indicate the detection limits. Dashed lines indicate the estimated viremia levels below the detection limits. Predicted mean viremia levels from 10th-root transformation are transformed to the log-10 scale. The viremia levels are shown for an age of 10 years and male sex.

### Appendix 6. Results for the effect of viremia on platelet count

**Appendix 6-table 1. P-values of parameters in the supermodel for platelet count**

| **Parameter** | **P value** | **Parameter** | **P value** |
| --- | --- | --- | --- |
| Age (overall effect) | <0.0001 | Age * Day | <0.0001 |
| - All interactions of Age | <0.0001 | Sex * Day | <0.0001 |
| - Nonlinear effect of Age | <0.0001 | Serotype * Day | <0.0001 |
| Sex (overall effect) | <0.0001 | Immune status * Day | <0.0001 |
| - All interactions of Sex | <0.0001 | PCR * Day | <0.0001 |
| Serotype (overall effect) | <0.0001 | Landmark * Day | <0.0001 |
| - All interactions of Serotype | <0.0001 | Viremia * Day | <0.0001 |
| Immune status (overall effect) | <0.0001 | Viremia * Serotype | <0.0001 |
| - All interactions of Immune status | <0.0001 | Viremia * Immune status | 0.0017 |
| PCR method (overall effect) | <0.0001 | Viremia * PCR | <0.0001 |
| - All interactions of PCR method | <0.0001 | Viremia * Landmark | <0.0001 |
| Day (overall effect) | <0.0001 | Serotype * Landmark | 0.0403 |
| - All interactions of Day | <0.0001 | PCR * Landmark | <0.0001 |
| - Nonlinear effect of Day | <0.0001 | Serotype * PCR | <0.0001 |
| Landmark (overall effect) | <0.0001 | Serotype * Immune status | <0.0001 |
| - All interactions of Landmark | <0.0001 | Viremia * Serotype * PCR | 0.2465 |
| - Nonlinear effect of Landmark | <0.0001 | Viremia * Serotype * Day | <0.0001 |
| Viremia (overall effect) | <0.0001 | Viremia * PCR * Day | <0.0001 |
| - All interactions of Viremia | <0.0001 | Viremia * Serotype * Landmark | 0.2601 |
| - Nonlinear effect of Viremia | <0.0001 | Viremia * PCR * Landmark | 0.0846 |
| - Undetectable Viremia | 0.1632 | Viremia * Serotype * Immune status | 0.0589 |
|  |  | Serotype * PCR * Day | <0.0001 |
|  |  | Serotype * PCR * Landmark | 0.1347 |
|  |  | Viremia * Serotype * PCR * Day | 0.4683 |
|  |  | Viremia * Serotype * PCR * Landmark | 0.3646 |

P values are calculated using the Wald test from the R package ‘aod’. This test computes the Wald *chi-squared* test for 1 or more coefficients given their variance-covariance matrix from the model. Note that the overall effect of viremia includes the binary variable describing detectable or undetectable viremia, and the term ‘Viremia’ in all interactions also consists of that binary variable.

A * B is the interaction between A and B.

PCR, polymerase chain reaction

**Appendix 6-figure 1. Fitted trends in mean platelet counts according to viremia levels by subgroups of the covariates - results from the supermodel**

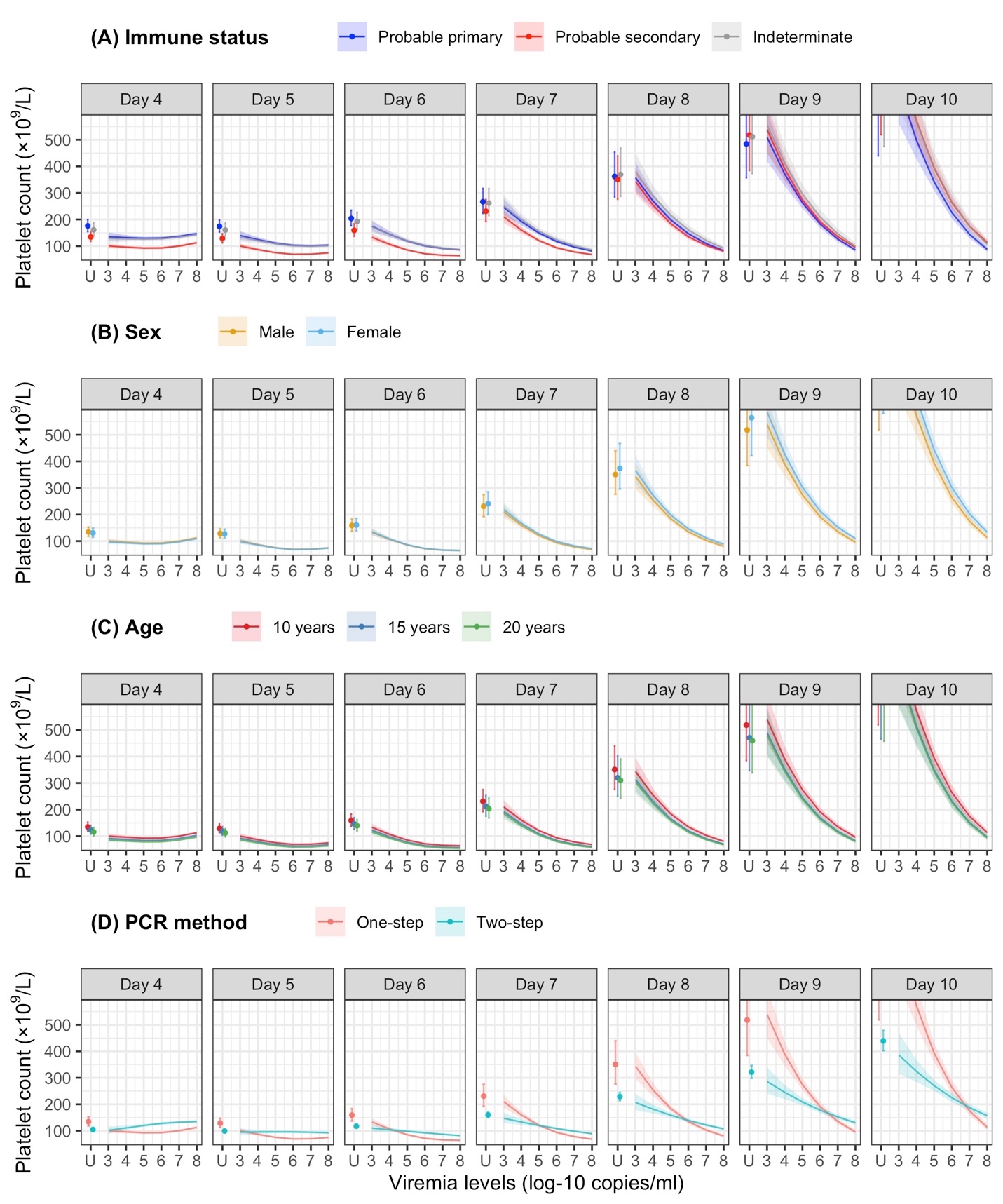

The colored lines or dots represent the estimated mean platelet counts. The colored shaded regions and whiskers indicate the corresponding 95% confidence intervals. The figures show the effect of viremia on day 4 to platelet count from day 4 to day 10. The mean platelet counts are shown for age of 10 years, male sex, serotype DENV-1, probable secondary infection, and using the one-step PCR. DENV, dengue virus; LM, landmark; PCR, polymerase chain reaction; U, under the limit of detection.

**Appendix 7. Results for the effect of viremia level on clinical outcomes**

### Appendix 7-table 1. Estimates from models of viremia and clinical outcomes

|  | **Severe dengue** | | **Plasma leakage** | |
| --- | --- | --- | --- | --- |
|  | **OR (95% CI)** | **p** | **OR (95% CI)** | **p** |
| Viremia level (log-10 copies/ml) ^a^ |  | <0.001 |  | <0.001 |
| - 7 versus 6 (on day 1) | 2.98 (1.46; 6.10) |  | 1.94 (1.50; 2.51) |  |
| - 8 versus 7 (on day 1) | 2.27 (1.20; 4.31) |  | 1.83 (1.44; 2.32) |  |
| - 7 versus 6 (on day 2) | 2.10 (1.32; 3.36) |  | 1.62 (1.36; 1.94) |  |
| - 8 versus 7 (on day 2) | 1.60 (1.05; 2.44) |  | 1.53 (1.29; 1.82) |  |
| - 7 versus 6 (on day 3) | 1.51 (1.13; 2.00) |  | 1.38 (1.20; 1.57) |  |
| - 8 versus 7 (on day 3) | 1.15 (0.84; 1.57) |  | 1.30 (1.11; 1.52) |  |
| - 7 versus 6 (on day 4) | 1.18 (0.91; 1.54) |  | 1.26 (1.10; 1.45) |  |
| - 8 versus 7 (on day 4) | 0.90 (0.61; 1.32) |  | 1.19 (0.98; 1.44) |  |
| - 7 versus 6 (on day 5) | 1.08 (0.75; 1.54) |  | 1.31 (1.10; 1.57) |  |
| - 8 versus 7 (on day 5) | 0.82 (0.49; 1.37) |  | 1.24 (0.96; 1.59) |  |
| Undetectable viremia (Yes vs. 3 log-10 copies/ml) | 0.48 (0.20; 1.16) | 0.102 | 1.84 (0.51; 6.65) | 0.247 |
| Age (years) ^a^ |  | 0.182 |  | 0.105 |
| - 15 versus 10 | 0.77 (0.50; 1.18) |  | 1.07 (0.88; 1.31) |  |
| - 20 versus 15 | 0.67 (0.44; 1.04) |  | 0.95 (0.85; 1.06) |  |
| Sex (Male vs. Female) | 1.43 (0.79; 2.61) | 0.011 | 1.10 (0.84; 1.45) | 0.711 |
| Serotype |  | <0.001 |  | <0.001 |
| - DENV-1 | Ref |  | Ref |  |
| - DENV-2 | 0.95 (0.44; 2.07) |  | 1.22 (0.84; 1.77) |  |
| - DENV-3 | 0.35 (0.08; 1.51) |  | 0.92 (0.54; 1.56) |  |
| - DENV-4 | 2.41 (0.83; 6.98) |  | 1.19 (0.76; 1.86) |  |
| Immune status |  | <0.001 |  | <0.001 |
| - Probable primary | 0.02 (0.00; 0.15) |  | 0.41 (0.27; 0.63) |  |
| - Probable secondary | Ref |  | Ref |  |
| - Indeterminate | 0.32 (0.04; 2.32) |  | 0.57 (0.30; 1.11) |  |
| Study |  | 0.010 |  | <0.001 |
| - Study C | Ref |  | Ref |  |
| - Study A | 1.36 (0.43; 4.28) |  | 1.24 (0.80; 1.93) |  |
| - Study B | 2.86 (1.21; 6.80) |  | 2.68 (1.92; 3.75) |  |
| Landmark (illness day) ^a^ |  | <0.001 |  | <0.001 |
| - 5 versus 3 | 1.38 (0.74; 2.57) |  | 1.37 (1.06; 1.78) |  |
| - 7 versus 5 | 0.58 (0.17; 2.05) |  | 1.34 (0.77; 2.35) |  |
| Interaction of viremia ^b^ |  |  |  |  |
| - With age |  | 0.237 |  | 0.044 |
| - With sex |  | 0.004 |  | 0.590 |
| - With landmark (illness day) |  | <0.001 |  | <0.001 |
| - With study |  | 0.030 |  | 0.261 |
| - With serotype |  | 1 |  | <0.001 |
| - With immune status |  | 1 |  | 0.005 |
| - With serotype * immune status |  | <0.001 |  | 0.023 |
| - With serotype * landmark |  | <0.001 |  | 0.014 |
| - With immune status * landmark |  | <0.001 |  | 0.016 |
| - With serotype * immune status * landmark |  | <0.001 |  | <0.001 |
| Interaction of serotype and immune status |  | <0.001 |  | <0.001 |
| Nonlinear trend |  |  |  |  |
| - Viremia level |  | 0.031 |  | 0.272 |
| - Age |  | 0.349 |  | 0.017 |
| - Landmark (illness day) |  | <0.001 |  | 0.048 |

^a^ The model incorporates nonlinear effects of log-10 viremia level, age, and illness day on the endpoints. To facilitate the interpretation of the results, ORs and their corresponding 95% CIs are provided for two selected contrasts of viremia level, age, and landmark illness day.

^b^ In the model for plasma leakage, the interactions of viremia include the binary variable describing detectable or undetectable viremia.

Given the complexity of the models involving multiple interactions, ORs and 95% CIs are estimated for specific values of the interacting variables: log-10 viremia of 7, age of 10 years, male sex, landmark illness day of 3, serotype DENV-1, and probable secondary infection. CI, confidence interval; DENV, dengue virus; OR, odds ratio; Ref, reference.

**Appendix 7-figure 1. Frequency of the outcomes and regression coefficients for individuals observed on each landmark day that did not have the outcome yet**

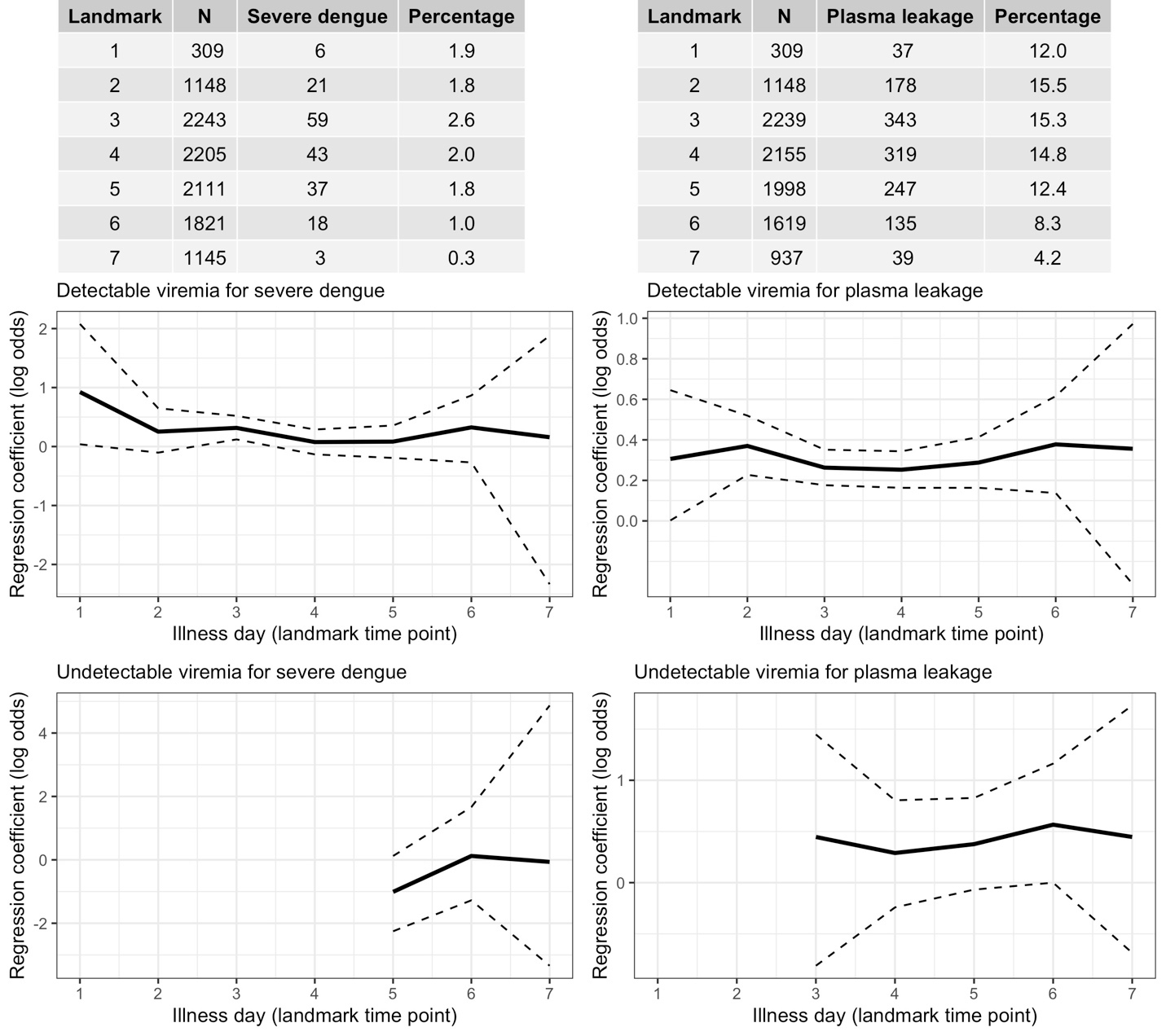

Relationship between viremia and outcome. A logistic regression model with covariates viremia level (with linear trend) and the binary variable describing whether the viremia value was undetectable is fitted per landmark time point. Solid lines are the regression coefficients and dashed lines are the 95% confidence intervals.

### Appendix 7-figure 2. Probability of occurrence of the two clinical endpoints according to viremia levels by subgroups of the covariates - results from the supermodel

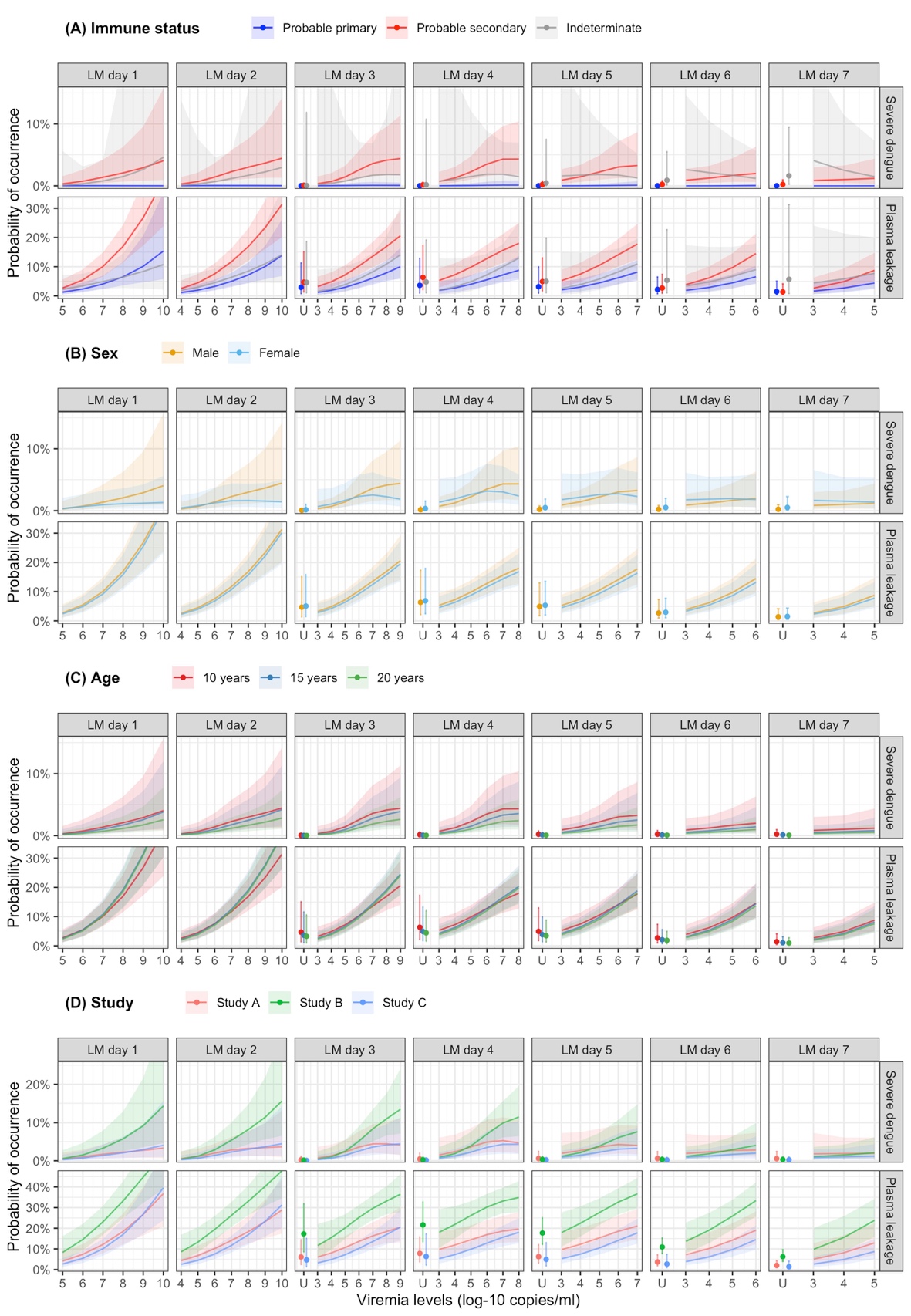

The colored lines or dots represent the probability of the endpoints. The colored shaded regions and whiskers indicate the corresponding 95% confidence intervals. Each column represents the effect of viremia on a specific day. Note that the probability of severe dengue in the probable primary group is almost 0% because there was only one case with severe dengue in this group. The probabilities are shown for age of 10 years, male sex, serotype DENV-1, probable secondary infection, and from Study C. LM, landmark; U, under the limit of detection.

### Appendix 7-Figure 3. Probability of occurrence of the two clinical endpoints according to viremia levels – compare results from the supermodels with and without multiple imputation

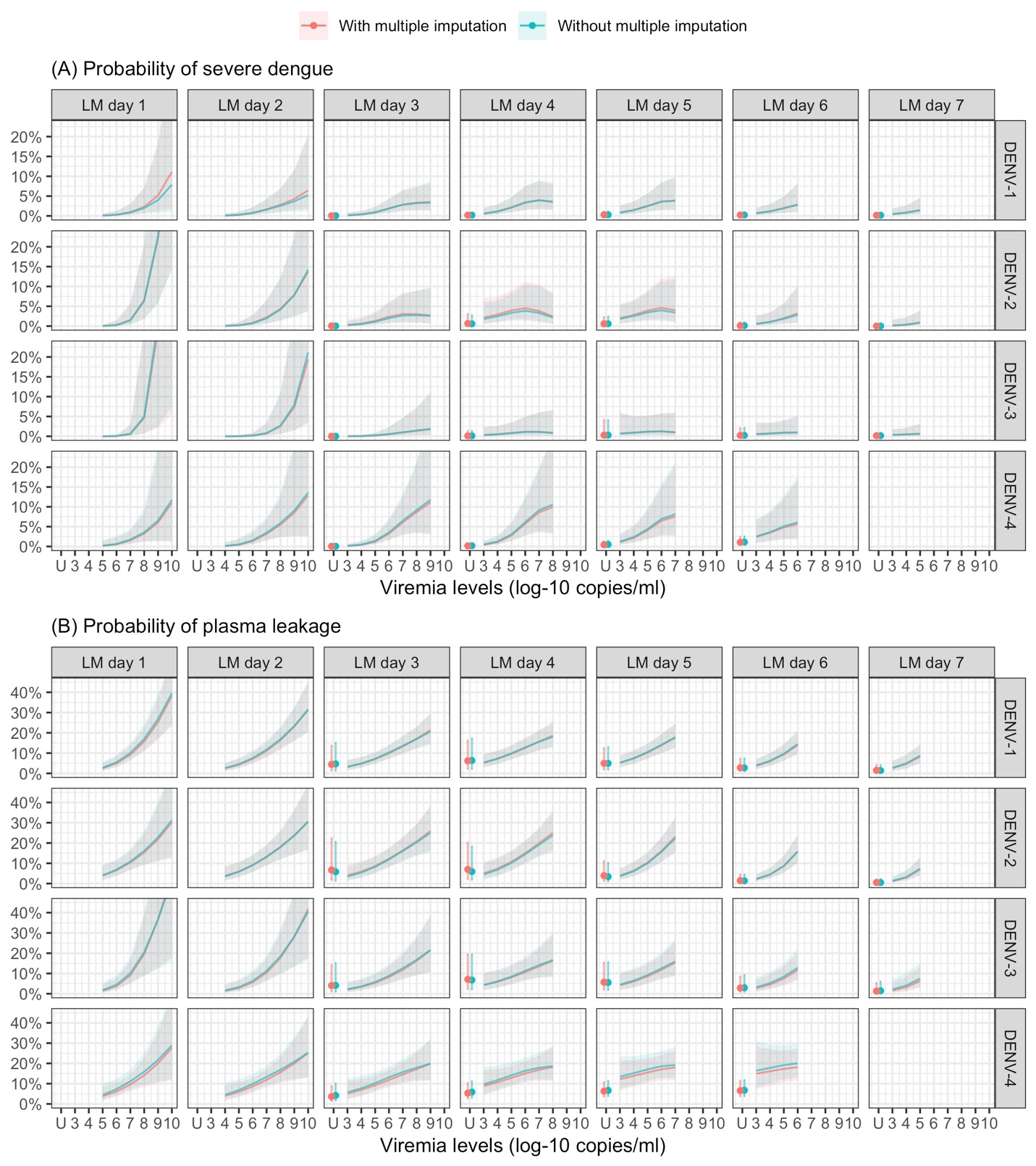

The colored lines or dots represent the probability of the endpoints. The colored shaded regions and whiskers indicate the corresponding 95% confidence intervals. Each column represents the effect of viremia on a specific day. No fitted trends are made for DENV-4 in LM day 7 since viremia was undetectable in almost all DENV-4 cases from day 7 onwards. The probabilities are shown for age of 10 years, male sex, probable secondary infection, and from Study C. DENV, dengue virus, LM, landmark; PCR, polymerase chain reaction; U, under the limit of detection.

**Appendix 7-figure 4. Probability of occurrence of the two clinical endpoints according to the covariates - results from the supermodel**

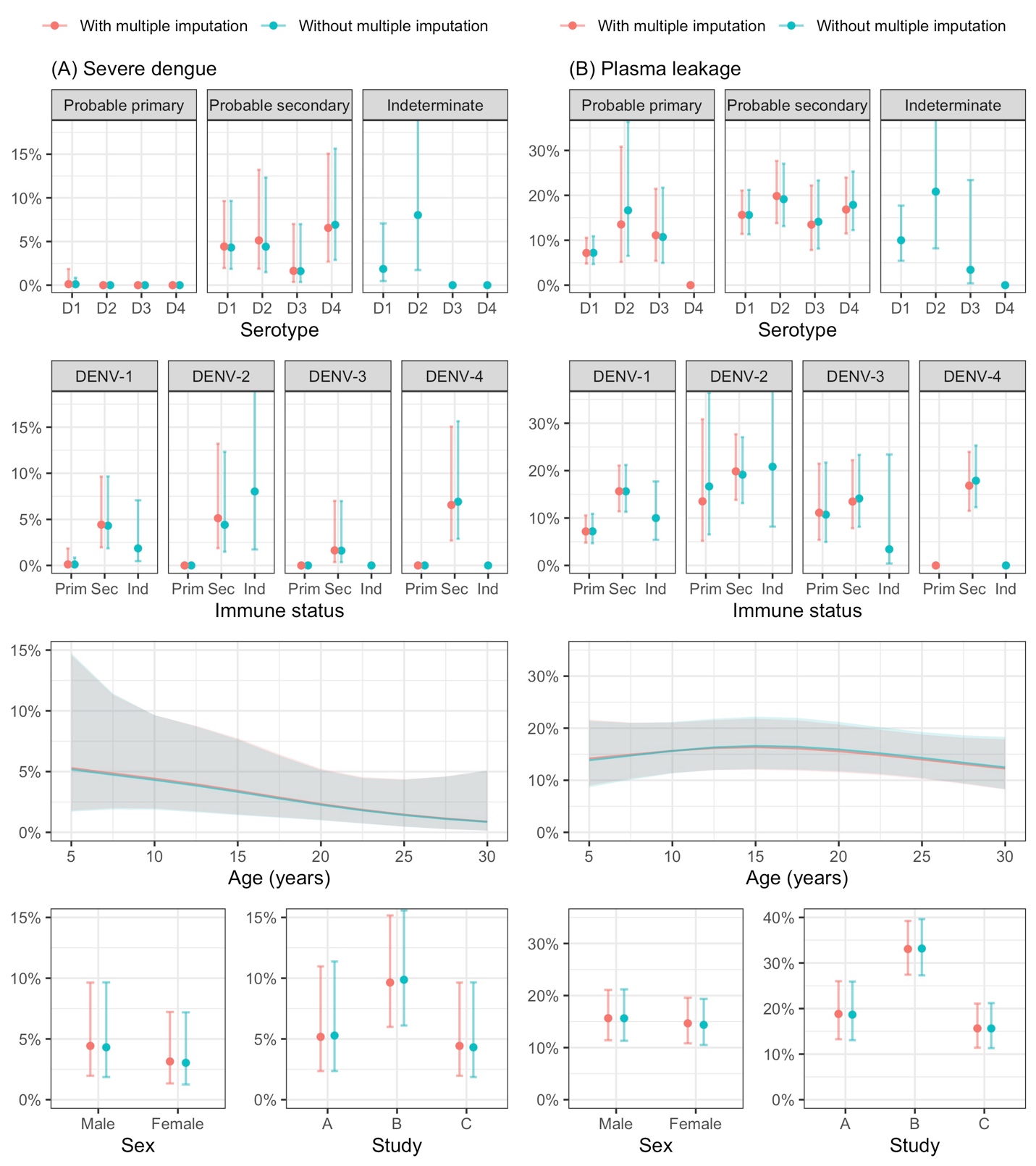

The lines and dots represent the probability of the endpoints. The shaded regions and whiskers indicate the corresponding 95% confidence intervals. Note that the probability of severe dengue in the probable primary group is almost 0% because there was only one case with severe dengue in this group. The probabilities are shown for age of 10 years, male sex, serotype DENV-1, probable secondary infection, viremia level of 7 log-10 copies/ml, landmark illness day 4, and from Study C. D1, D2, D3, and D4 are DENV-1, DENV-2, DENV-3, and DENV-4, respectively; Prim, probable primary infection; Sec, probable secondary infection; Ind, Indeterminate immune status.

### Appendix 7-table 2. Estimates from models of the rate of decline in viremia and clinical outcomes

|  | **Severe dengue** | | **Plasma leakage** | |
| --- | --- | --- | --- | --- |
|  | **OR (95% CI)** | **p** | **OR (95% CI)** | **p** |
| Rate of decline in viremia (per 0.5 log-10 copies/ml increase) | 0.14 (0.06; 0.33) | <0.001 |  | <0.001 |
| - DENV-1 - probable primary |  |  | 0.23 (0.09; 0.56) |  |
| - DENV-1 - probable secondary |  |  | 0.12 (0.05; 0.28) |  |
| - DENV-2 - probable primary |  |  | 0.30 (0.11; 0.80) |  |
| - DENV-2 - probable secondary |  |  | 0.16 (0.06; 0.38) |  |
| - DENV-3 - probable primary |  |  | 0.18 (0.06; 0.54) |  |
| - DENV-3 - probable secondary |  |  | 0.09 (0.03; 0.27) |  |
| - DENV-4 - probable primary |  |  | 0.78 (0.24; 2.50) |  |
| - DENV-4 - probable secondary |  |  | 0.40 (0.16; 1.04) |  |
| Serotype |  | 0.001 |  | 0.006 |
| - DENV-1 | Ref |  | Ref |  |
| - DENV-2 | 0.74 (0.39; 1.39) |  | 1.04 (0.75; 1.45) |  |
| - DENV-3 | 0.21 (0.05; 0.88) |  | 0.81 (0.54; 1.22) |  |
| - DENV-4 | 8.59 (2.38; 31.03) |  | 4.84 (2.07; 11.32) |  |
| Immune status |  | <0.001 |  | <0.001 |
| - Probable primary | Ref |  | Ref |  |
| - Probable secondary | 63.23 (8.15; 490.31) |  | 2.77 (1.78; 4.33) |  |
| - Indeterminate | 16.29 (1.77; 150.25) |  | 1.39 (0.68; 2.82) |  |
| PCR method |  | <0.001 |  | <0.001 |
| - One-step | Ref |  | Ref |  |
| - Two-step | 33.32 (10.92; 101.66) |  | 14.62 (6.98; 30.60) |  |
| Interaction of rate of decline in viremia |  |  |  | 0.056 |
| - With serotype |  |  |  | 0.182 |
| - With immune status |  |  |  | 0.017 |
| - With PCR |  |  |  | 0.006 |

The model for severe dengue does not include any interaction term.

In the model of plasma leakage, interactions between the rate of decline in viremia and serotype, immune status, and PCR are included. ORs and 95% CIs of the rate of decline in viremia are estimated for the one-step PCR in each subgroup of serotype and probable primary/secondary infection. ORs and 95% CIs of serotype, immune status, and PCR are estimated for the rate of decline in viremia of 1.4 log-10 copies/ml.

CI, confidence interval; DENV, dengue virus; OR, odds ratio; PCR, polymerase chain reaction; Ref, reference

**Appendix 8. STROBE Statement - Checklist of items**

**Dengue viremia kinetics and relationship with platelet count and clinical outcomes: an analysis of 2340 patients from Vietnam**

|  | **Item No** | **Recommendation** | **Comments or page(s) reported** |
| --- | --- | --- | --- |
| **Title and abstract** | 1 | (*a*) Indicate the study’s design with a commonly used term in the title or the abstract | Page 1 |
|  |  | (*b*) Provide in the abstract an informative and balanced summary of what was done and what was found | Page 2 |
| **Introduction** | | |  |
| Background/rationale | 2 | Explain the scientific background and rationale for the investigation being reported | Pages 3-4 |
| Objectives | 3 | State specific objectives, including any prespecified hypotheses | Page 4 |
| **Methods** | | |  |
| Study design | 4 | Present key elements of study design early in the paper | Pages 4-5 |
| Setting | 5 | Describe the setting, locations, and relevant dates, including periods of recruitment, exposure, follow-up, and data collection | Pages 4-5, Appendix 1 |
| Participants | 6 | (*a*) Give the eligibility criteria, and the sources and methods of selection of participants. Describe methods of follow-up | Pages 4-5, Appendix 1 |
|  |  | (*b*) For matched studies, give matching criteria and number of exposed and unexposed | Not applicable |
| Variables | 7 | Clearly define all outcomes, exposures, predictors, potential confounders, and effect modifiers. Give diagnostic criteria, if applicable | Page 5, Appendix 2 |
| Data sources/ measurement | 8* | For each variable of interest, give sources of data and details of methods of assessment (measurement). Describe comparability of assessment methods if there is more than one group | Page 5 |
| Bias | 9 | Describe any efforts to address potential sources of bias | Pages 5-7 |
| Study size | 10 | Explain how the study size was arrived at | Pages 5-7, Figure 1 |
| Quantitative variables | 11 | Explain how quantitative variables were handled in the analyses. If applicable, describe which groupings were chosen and why | Pages 6-7, Appendix 3 |
| Statistical methods | 12 | (*a*) Describe all statistical methods, including those used to control for confounding | Pages 6-7, Appendix 3 |
|  |  | (*b*) Describe any methods used to examine subgroups and interactions | Pages 6-7, Appendix 3 |
|  |  | (*c*) Explain how missing data were addressed | Page 7, Appendix 3 |
|  |  | (*d*) If applicable, explain how loss to follow-up was addressed | Not available |
|  |  | (*e*) Describe any sensitivity analyses | Pages 6-7, Appendix 3 |
| **Results** | | |  |
| Participants | 13* | (a) Report numbers of individuals at each stage of study—eg numbers potentially eligible, examined for eligibility, confirmed eligible, included in the study, completing follow-up, and analysed | Pages 5-6, Figure 1 |
|  |  | (b) Give reasons for non-participation at each stage | Pages 5-6, Figure 1 |
|  |  | (c) Consider use of a flow diagram | Figure 1 |
| Descriptive data | 14* | (a) Give characteristics of study participants (eg demographic, clinical, social) and information on exposures and potential confounders | Pages 7-8, Table 1 |
|  |  | (b) Indicate number of participants with missing data for each variable of interest | Pages 7-8, Table 1 |
|  |  | (c) Summarise follow-up time (eg, average and total amount) | Not available |
| Outcome data | 15* | Report numbers of outcome events or summary measures over time | Pages 7-8, Table 1, Appendix 4-table 1 |
| Main results | 16 | (*a*) Give unadjusted estimates and, if applicable, confounder-adjusted estimates and their precision (eg, 95% confidence interval). Make clear which confounders were adjusted for and why they were included | Pages 8-9,  Appendices 5-7 |
|  |  | (*b*) Report category boundaries when continuous variables were categorized | Not available |
|  |  | (*c*) If relevant, consider translating estimates of relative risk into absolute risk for a meaningful time period | Pages 8-9,  Appendices 5-7 |
| Other analyses | 17 | Report other analyses done—eg analyses of subgroups and interactions, and sensitivity analyses | Pages 8-9,  Appendices 5-7 |
| **Discussion** | | |  |
| Key results | 18 | Summarise key results with reference to study objectives | Page 10 |
| Limitations | 19 | Discuss limitations of the study, taking into account sources of potential bias or imprecision. Discuss both direction and magnitude of any potential bias | Page 12 |
| Interpretation | 20 | Give a cautious overall interpretation of results considering objectives, limitations, multiplicity of analyses, results from similar studies, and other relevant evidence | Pages 10-12 |
| Generalisability | 21 | Discuss the generalisability (external validity) of the study results | Pages 11-12 |
| **Other information** | | |  |
| Funding | 22 | Give the source of funding and the role of the funders for the present study and, if applicable, for the original study on which the present article is based | Page 14 |

*Give information separately for exposed and unexposed groups.

**Note:** An Explanation and Elaboration article discusses each checklist item and gives methodological background and published examples of transparent reporting. The STROBE checklist is best used in conjunction with this article (freely available on the Web sites of PLoS Medicine at http://www.plosmedicine.org/, Annals of Internal Medicine at http://www.annals.org/, and Epidemiology at http://www.epidem.com/). Information on the STROBE Initiative is available at http://www.strobe-statement.org.
